# Upper respiratory microbiome and host immune signatures associated with SARS-CoV-2 susceptibility and illness in a human challenge model

**DOI:** 10.64898/2026.11.16.26368138

**Authors:** Jillian H. Hurst, Visanu Wanchai, Pixu Shi, Intawat Nookaew, Devjanee Swain-Lenz, Laura-Leigh Rowlette, Sarah Cleaton, Jason Arnold, So Young Kim, Corbin Hite, Kristine L. Kay, Andrew Catchpole, Mariya Kalinova, Ben Killingley, Alex Mann, Helen Wagstaffe, Christopher Chiu, Matthew S. Kelly

## Abstract

The upper respiratory tract (URT) microbiome modulates the local mucosal immune environment, with potential implications for respiratory viral infection susceptibility and outcomes. We leveraged a first-in-human SARS-CoV-2 challenge study in 34 healthy young adults, 18 of whom became infected, to characterize host gene expression and microbiome composition and dynamics in nasal and throat samples collected before viral inoculation and over the subsequent 14 days. Compared with participants who remained uninfected, participants who became infected had elevated baseline nasal expression of gene modules related to interferon, innate inflammatory, and neutrophil responses, as well as a delayed transcriptional response to viral challenge. Baseline nasal microbiome composition differed modestly by subsequent infection status, with lower relative abundances of *Corynebacterium accolens* and higher abundances of *Dolosigranulum pigrum* and *Staphylococcus aureus* among participants who became infected. Integrated analyses of microbiome and host gene expression data identified coordinated networks of microbiome features and host immune modules that distinguished infected from uninfected participants, including prior to inoculation. Notably, immune modules associated with infection susceptibility were positively correlated with *S. aureus* abundance and negatively correlated with the abundances of common nasal commensals, *Corynebacterium* spp. and *Staphylococcus epidermidis*, linking altered nasal microbiome community structure with the heightened inflammatory state associated with infection susceptibility. Symptom burden and viral load were similarly linked to distinct networks of microbiome and host gene expression features within nasal samples. Taken together, these findings highlight potential microbiome–host interactions within the nasal mucosa that may contribute to differences in respiratory viral infection outcomes.

## One Sentence Summary

Analyses from a first-in-human SARS-CoV-2 challenge study identified microbiome–host immune networks associated with infection susceptibility and outcomes.

## INTRODUCTION

The mucosa of the upper respiratory tract (URT) serves as the first line of defense against respiratory pathogens (*1*). This highly specialized epithelium provides a physical barrier that traps and removes pathogens while limiting their invasion beyond the URT. Epithelial cells within the URT also initiate innate immune responses that can directly prevent colonization or infection and recruit immune cells from the periphery to promote pathogen clearance (*2*). Thus, the URT epithelium plays a central role in determining susceptibility to respiratory infection and shaping the host responses that contribute to the symptoms of established infection.

Accumulating evidence suggests that the structure and function of the URT epithelium are closely linked to the resident microbial communities within this niche. These microbes interact with the host and with one another, forming a complex ecosystem that can protect against pathogen colonization and invasion (*3*). Previous studies have shown that the presence and abundance of specific microbes in the URT can influence the risk and outcomes of respiratory viral infections, primarily in the context of influenza (*4, 5*). For example, colonization by bacterial pathobionts such as *Haemophilus influenzae* and *Streptococcus pneumoniae* has been shown to increase the severity of respiratory viral infections by altering epithelial and immune responses and directly promoting viral replication, adherence, and entry (*6–10*). Together, these findings demonstrate an important role for the microbiome in influencing host defense in the URT and respiratory viral pathobiology.

We and others previously showed that the URT microbiome is associated with SARS-CoV-2 infection susceptibility and symptom burden; however, the relationships between specific microbes and host immune responses that may underly these associations have not been defined (*11, 12*). Multiple studies also identified distinct URT host responses in SARS-CoV-2-infected individuals compared with those who were exposed but remained uninfected. However, these data have come primarily from cross-sectional studies that included participants who varied widely in age, baseline health status, stage of infection at sampling, and prior infection history (*13–15*). Human challenge studies, which involve the intentional infection of research participants with a standardized microbial inoculum under precisely timed and controlled conditions, can overcome many of these limitations inherent to natural infection studies. Herein, we leveraged clinical data and nasal and throat swab samples from a first-in-human SARS-CoV-2 challenge study to define host immune responses and microbial community dynamics within the URT and their relationships with infection outcomes (*16*). We performed integrative analyses of longitudinal microbiome and host gene expression data to identify microbe–host immune interactions associated with infection susceptibility, symptom burden, and viral kinetics.

## RESULTS

### Differential URT host responses in SARS-CoV-2-infected versus uninfected participants

Thirty-four healthy adult volunteers aged 18 to 29 years who were seronegative for SARS-CoV-2 anti-Spike antibodies were inoculated with a wild-type SARS-CoV-2 (D614G) challenge virus; 18 (53%) subsequently developed PCR-confirmed infection (*16*). To assess host gene expression in the URT before and after viral inoculation, we performed bulk RNA sequencing of mid-turbinate nasal and throat swab samples collected in viral transport medium (VTM) at a single pre-inoculation timepoint (−1am) and throughout the 14 days after inoculation. To characterize the expression of relevant immune pathways, we used a modular analytic approach that groups transcripts based on shared patterns of expression observed in other human infections and disease states (*17*). We selected 17 modules from this framework based on their levels of expression in this dataset and representation of key immune pathways involved in infection response (**Supplemental File 1**). Modular expression at the sample level was quantified using single-sample gene set enrichment analysis (ssGSEA).

We first compared URT gene expression between participants who developed PCR-confirmed infection and those who remained uninfected after viral challenge (**Fig. 1**). Prior to inoculation, overall nasal gene expression differed significantly by subsequent infection status (PERMANOVA, R²=0.098, p=0.043). Participants who acquired infection had higher nasal expression of modules related to interferon-stimulated gene responses (ISGs; M3.4), antiviral sensing (M5.4), innate immune signaling and inflammation (M4.13, M9.51), phagocytosis (M5.1), neutrophil responses (M4.9, M7.1), and toll-like receptor (TLR) signaling (M3.2) (**Fig. 1A**), whereas those who remained uninfected had higher expression of T cell-associated responses (M4.1). By the first day after viral inoculation, uninfected individuals exhibited broad changes in nasal immune responses relative to baseline, with increased expression of interferon, innate inflammatory, and neutrophil-associated responses, and decreased expression of B cell responses (**Fig. 1B**). Many of the immune modules that increased early after inoculation overlapped with modules that had higher expression at baseline among participants who subsequently became infected. In contrast, infected participants exhibited little change in nasal gene expression relative to baseline until day 3, when interferon responses (M1.2, M3.4) were upregulated, often coinciding with a rapid increase in viral load (**Fig. S1**). As infection progressed, this immune activation broadened to include upregulation of modules corresponding to monocyte/antigen-presenting cell (APC) activation (M4.6), neutrophil degranulation (M5.7), NK cells (M3.6), and B cells (M4.2) (**Fig. 1B**). Expression of many of these modules remained elevated through 14 days after viral inoculation, although interferon and antiviral sensing responses had declined substantially from their peaks on day 7. Direct comparison of immune module expression between infected and uninfected participants at each timepoint further highlighted these divergent longitudinal patterns, with differences in innate immune and inflammatory responses evident both before and after viral inoculation (**Fig. 1C**).

**Figure 1.**
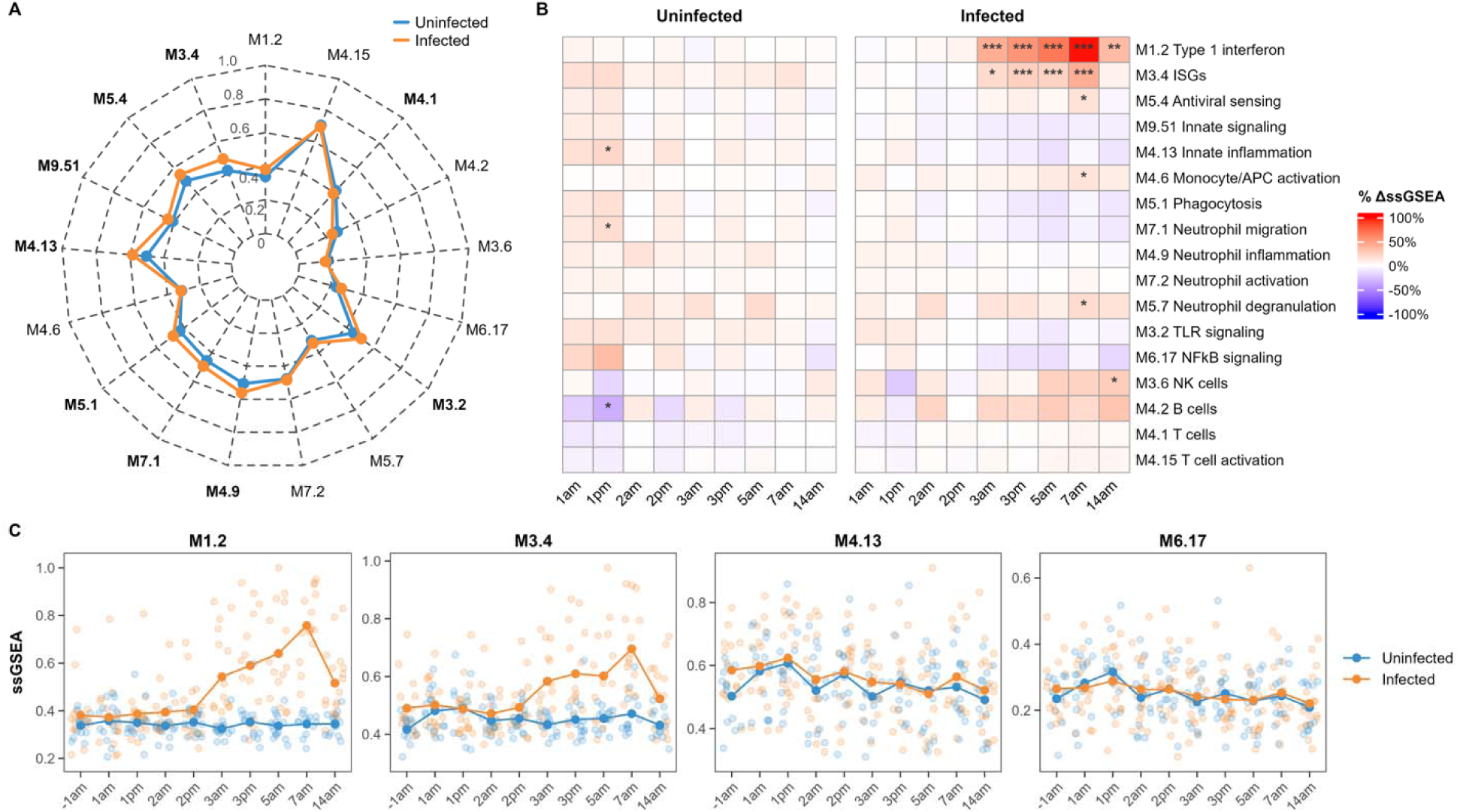
Nasal immune module expression associated with SARS-CoV-2 infection susceptibility. **(A)** Radar plot depicting baseline (−1am) nasal immune module expression in participants who remained uninfected (blue) or subsequently became infected (orange). Module expression was quantified using single-sample gene set enrichment analysis (ssGSEA) and scaled to a 0–1 range within the nasal dataset, where 0 and 1 correspond to the minimum and maximum observed values, respectively. Values represent group mean scaled ssGSEA scores. Module labels shown in bold indicate baseline differences between groups at a false discovery rate (FDR)-adjusted q-value < 0.20. **(B)** Heatmaps depicting longitudinal changes in nasal immune module expression relative to baseline within uninfected and infected participants. Values represent percent change from baseline mean scaled ssGSEA scores for each group. Statistical significance was determined using age- and sex-adjusted linear mixed-effects models including timepoint, infection status, and their interaction as fixed effects, with participant included as a random effect. Asterisks indicate FDR-adjusted significance for within-group changes relative to baseline, with FDR correction performed across modules at each timepoint within each infection group (* q < 0.20, ** q < 0.05, *** q < 0.01). **(C)** Longitudinal trajectories of the expression of selected immune modules in nasal samples by infection status. Points represent individual participant observations, while lines and larger points represent group mean scaled ssGSEA values at each timepoint.

In contrast to the nasal cavity, gene expression in the throat did not differ by subsequent infection status prior to inoculation (PERMANOVA, R²=0.023, p=0.58, **Fig. S2A**). During the first 2 days following challenge, participants who subsequently became infected and those who remained uninfected exhibited little change in throat gene expression relative to baseline (**Fig. S2B**), although NF-κB signaling (M6.17) and NK cell (M3.6) responses were generally upregulated in uninfected participants and remained elevated through day 14. By day 3, infected participants exhibited broad activation of innate immune modules, including those for interferon signaling, innate inflammation, phagocytosis, and neutrophil responses, with these responses largely returning to baseline by day 14. Direct comparison of infected and uninfected participants similarly showed little difference in throat gene expression during the first 2 days following viral inoculation (**Fig. S2C**). Thus, throat immune responses were relatively muted early after viral exposure, with broader changes emerging by day 3 in participants who became infected.

### URT microbiome composition in SARS-CoV-2-infected and uninfected participants

We characterized URT microbiome composition using 16S rRNA gene and shotgun metagenomic sequencing of nasal and throat samples, respectively, and quantified bacterial load using a qPCR assay targeting the 16S rRNA gene. Alpha diversity, richness, and bacterial load did not differ across timepoints by infection status in either sample type (**Fig. S3**). At baseline, nasal microbiome composition differed modestly by subsequent infection status (PERMANOVA, R²=0.065, p=0.08), whereas throat microbiome composition was similar in these groups (PERMANOVA, R²=0.027, p=0.42). Participants who subsequently became infected tended to have lower relative abundances of *Corynebacterium accolens* and higher relative abundances of *Dolosigranulum pigrum* and *Staphylococcus aureus* (**Fig. 2A**). In contrast, baseline relative abundances of microbial species in throat samples were remarkably similar between participants who subsequently became infected and those who remained uninfected (**Fig. S4A**).

**Figure 2.**
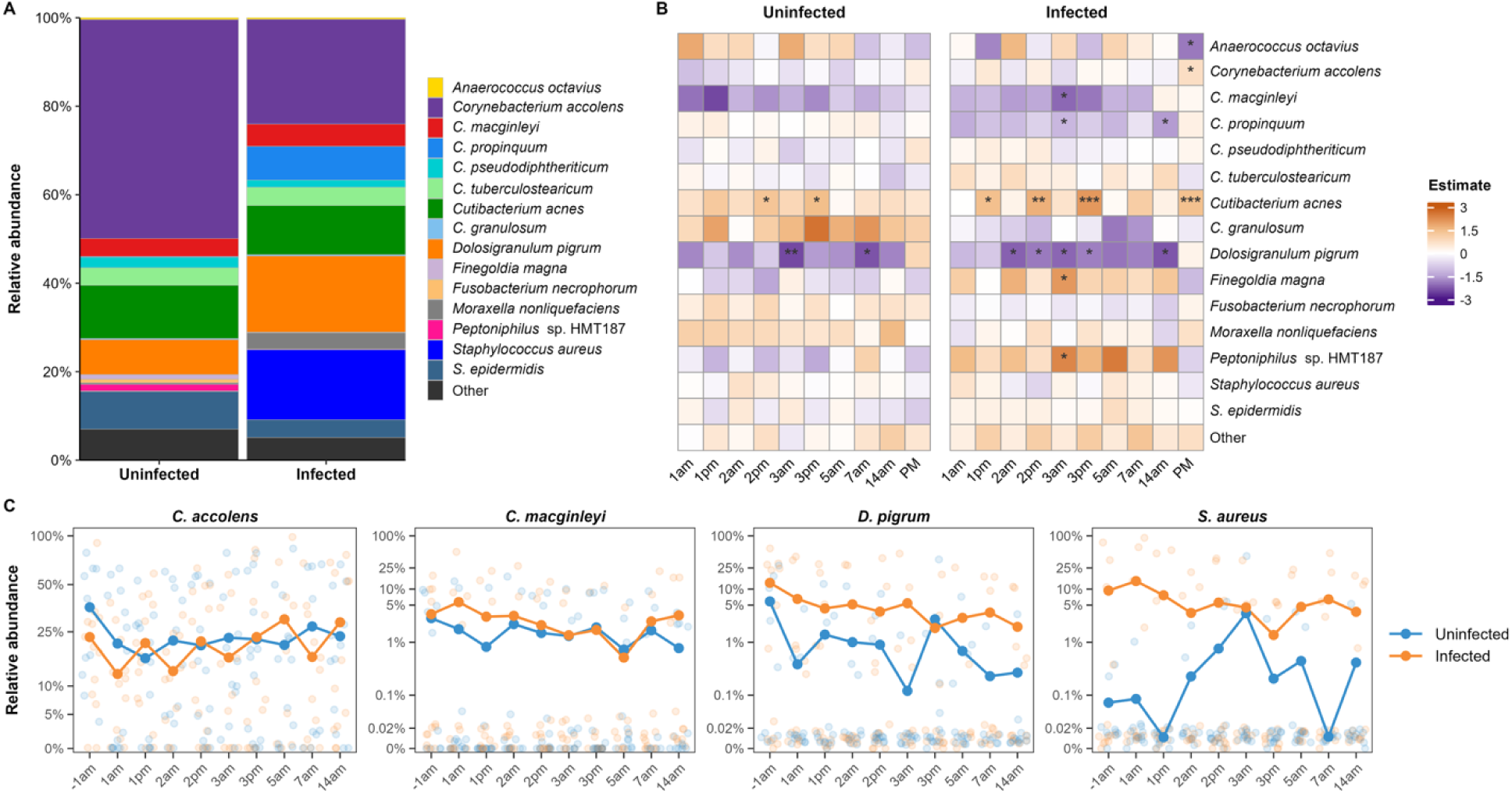
Nasal microbiome features associated with SARS-CoV-2 infection. **(A)** Stacked bar plots depicting baseline (−1am) nasal microbiome composition in participants who remained uninfected (blue) or subsequently became infected (orange). Values represent mean relative abundances of retained bacterial species, with remaining species grouped as “Other.” No individual species differed significantly in baseline relative abundance between groups after correction for multiple comparisons. **(B)** Heatmaps depicting longitudinal changes in nasal species relative abundance compared to baseline within uninfected and infected participants. Values represent model-estimated changes in centered log-ratio (CLR)-transformed abundance derived from age- and sex-adjusted linear mixed-effects models incorporating day, time of day, infection status, their interactions, and participant-level random effects. Asterisks indicate false discovery rate (FDR)-adjusted significance for within-group changes relative to baseline (* q<0.20, ** q<0.05, *** q<0.01). **(C)** Longitudinal trajectories of selected nasal bacterial species by infection status. Points represent individual participant observations, while lines and larger points represent group mean relative abundances at each timepoint. Relative abundance is displayed on a pseudo-logarithmic scale to facilitate visualization of low-abundance species.

To evaluate whether microbiome community composition changed after viral inoculation, we used linear mixed-effects models to estimate changes in the abundance of individual species at each timepoint relative to baseline within infected and uninfected participants (**Fig. 2B, Fig. S4B**). In nasal samples, relatively few changes in species abundance were observed after viral inoculation, regardless of infection status. In infected participants, *Cutibacterium acnes* was enriched and *D. pigrum* was depleted at several post-inoculation timepoints relative to baseline, with similar but somewhat less pronounced changes observed in uninfected participants (**Fig 2B**– **C**). By comparison, the throat microbiome showed more extensive changes following viral inoculation, although the patterns of change were broadly similar between infected and uninfected participants (**Fig. S4B**–**S4C**). A notable exception was non-meningococcal *Neisseria* spp., several of which became progressively enriched following inoculation in uninfected participants but not in those who became infected.

Superimposed on these longitudinal changes in taxon abundances after viral inoculation, we identified distinct diurnal patterns in multiple species (**Fig. 2B, Fig. S4B**). These patterns were more pronounced in throat samples, where the abundances of a substantial proportion of species differed between morning and afternoon samples. In both infected and uninfected participants, *Gemella haemolysans* and *Haemophilus parahaemolyticus* were more abundant in afternoon than morning throat samples, whereas *Prevotella* spp. and *Veillonella* spp. were less abundant (**Fig. S4B**). Additional diurnal shifts in species abundances were observed among infected participants, including higher abundances of several *Streptococcus* spp. in afternoon samples, although these patterns did not differ significantly by infection status.

### Multi-omic signatures associated with SARS-CoV-2 infection

We next sought to identify coordinated microbiome and host immune signatures associated with SARS-CoV-2 infection susceptibility. We used DIABLO, a supervised multi-omics integration method, to analyze URT microbiome and host immune data together (*18*). DIABLO constructs latent components that identify correlations between features across omics datasets while also distinguishing between prespecified groups. Separate models were constructed for nasal and throat samples at each timepoint to identify microbial species and immune modules that were correlated with each other and together distinguished infected from uninfected participants. In baseline nasal samples, the DIABLO model had an area under the receiver operating characteristic curve (AUC) of 0.81 for immune module data and 0.78 for microbiome data, indicating moderate discrimination between participants who subsequently became infected and those who remained uninfected (**Fig. 3A**). Classification accuracy was 65% at baseline, indicating that microbiome and immune module data collected before viral inoculation could correctly classify the subsequent infection status of approximately two-thirds of participants. Both the AUC and classification accuracy of the nasal models began to increase on day 3, with discrimination improving from morning to afternoon and at subsequent timepoints, driven primarily by immune module data. Throat models showed weaker discrimination at baseline than nasal models (**Fig. S5A**), with AUCs <0.70 for both microbiome and immune module data and a classification accuracy of 55%. Throat models showed a similar increase in discriminatory performance at later timepoints following the establishment of infection.

**Figure 3.**
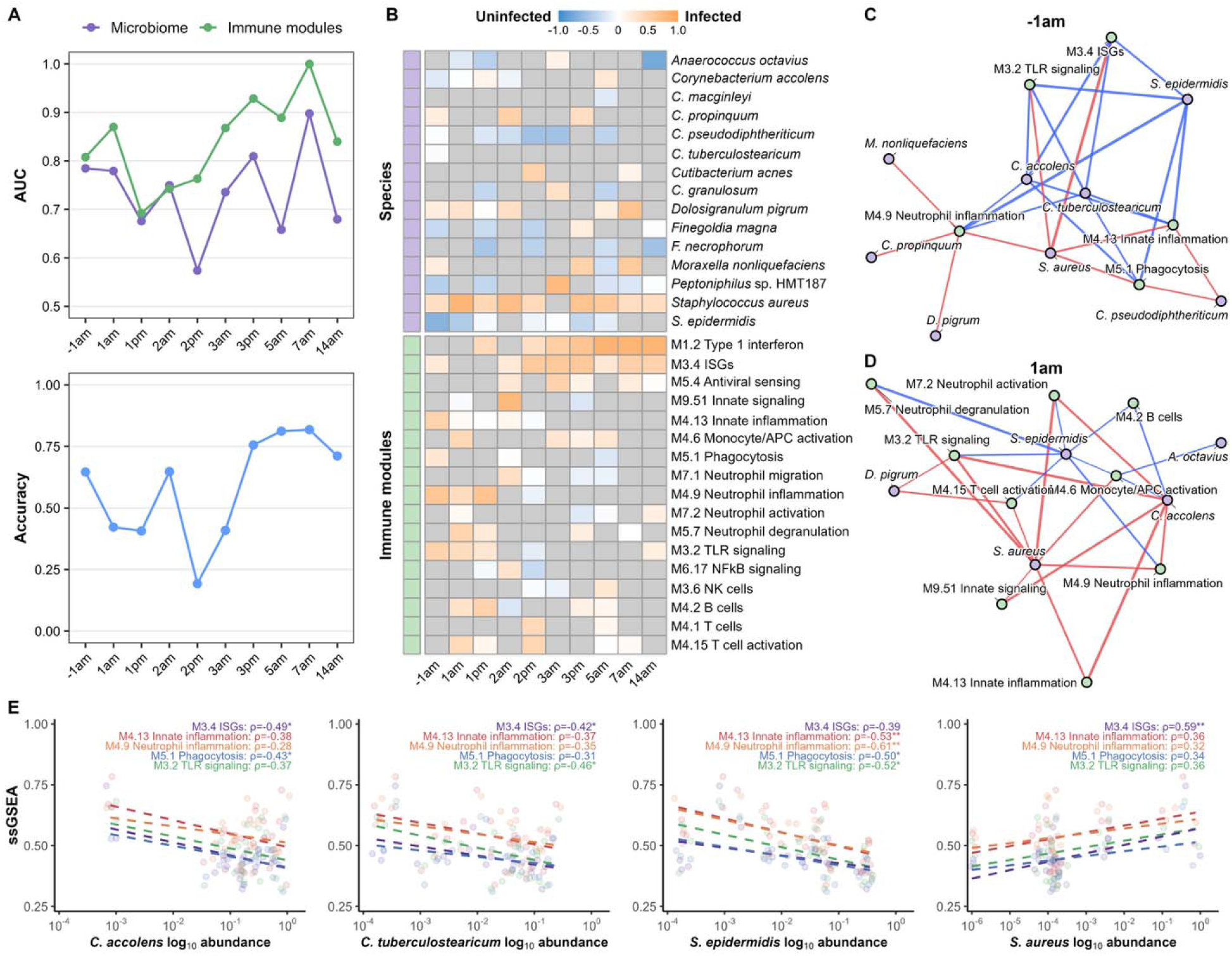
Nasal microbiome-host immune signatures associated with SARS-CoV-2 infection susceptibility. **(A)** Performance of DIABLO models trained to distinguish infected and uninfected participants at each study timepoint using nasal bacterial species abundance and host immune module expression data. The upper panel shows component 1 area under the receiver operating characteristic curve (AUC) for microbiome and immune-module data blocks. The lower panel shows overall classification accuracy based on Mahalanobis distance. **(B)** Heatmap of signed component 1 loadings from DIABLO models across longitudinal timepoints. Loadings were oriented such that positive values indicate features associated with infected participants and negative values indicate features associated with uninfected participants. Species and immune modules selected by DIABLO are shown. **(C, D)** Microbe–host immune networks at baseline (− 1am) and 1am. Networks were constructed using DIABLO-selected microbial species and immune modules. Edges represent Pearson correlations between log_10_-transformed microbial relative abundance and scaled ssGSEA immune-module scores, with edge color indicating positive (red) or negative (blue) associations and edge width indicating correlation strength. To enhance interpretability, only the 25 strongest correlations at each timepoint are displayed. **(E)** Associations between selected nasal bacterial species and host immune modules at baseline. Points represent individual participants. Dashed lines represent linear regression fits. Correlation coefficients (ρ) and nominal p values are shown for each immune module.

We first evaluated which specific microbiome and host immune features contributed to discrimination between infected and uninfected participants over time. DIABLO generates loadings for features in each omics dataset that discriminate between prespecified groups, in this case participants who became infected and those who remained uninfected (**Fig. 3B, Fig. S5B**). In baseline (−1am) nasal samples, immune modules for ISGs (M3.4), innate inflammation (M4.13), phagocytosis (M5.1), neutrophil inflammation (M4.9), and TLR signaling (M3.2) were positively associated with subsequent infection, consistent with our prior finding that participants who became infected had elevated baseline expression of several of these modules (**Fig. 1A**). The morning after viral inoculation (1am), we observed continued associations between subsequent infection status and the expression of modules for innate immune and inflammatory responses (M4.13, M9.51), monocyte/APC activation (M4.6), neutrophil responses (M4.9, M5.7, M7.2), TLR signaling (M3.2), B cells (M4.2), and T cell activation (M4.15). Microbiome features also distinguished infected and uninfected participants, with higher baseline abundances of *Corynebacterium propinquum*, *D. pigrum*, *Moraxella nonliquefaciens*, and *S. aureus* associated with subsequent infection, and higher abundances of *C. accolens*, *Corynebacterium pseudodiphtheriticum*, *Corynebacterium tuberculostearicum*, *Finegoldia magna*, *Peptoniphilus sp.* HMT187, and *Staphylococcus epidermidis* associated with remaining uninfected (**Fig. 3B**). Despite weaker overall performance, the baseline throat model selected generally similar immune features, with higher expression of innate immune and inflammatory modules associated with subsequent infection (**Fig. S5B**). Among throat microbiome features, higher baseline abundances of *G. haemolysans*, *Streptococcus oris*, and *S. pneumoniae* were associated with subsequent infection, whereas higher abundances of *Prevotella melaninogenica* and *Veillonella nakazawae* were associated with remaining uninfected.

We next examined relationships between microbiome and host immune features associated with infection status by constructing correlation networks among the bacterial species and immune modules selected by DIABLO at each timepoint. At baseline (−1am; **Fig. 3C**), features associated with subsequent infection formed a coordinated network, with modules for ISGs (M3.4), TLR signaling (M3.2), innate inflammation (M4.13), neutrophil inflammation (M4.9), and phagocytosis (M5.1) positively correlated with the relative abundances of *C. propinquum*, and *D. pigrum, M. nonliquefaciens*, and *S. aureus*. In contrast, the relative abundances of *C. accolens*, *C. tuberculostearicum*, and *S. epidermidis*, which were associated with remaining uninfected, were negatively correlated with these same immune modules. Many of these relationships remained evident the morning after viral inoculation (1am; **Fig. 3D**), with *D. pigrum* and *S. aureus* positively correlated with innate and inflammatory immune modules, whereas *C. accolens* and *S. epidermidis* showed predominantly negative relationships. Selected relationships from the baseline correlation network were further examined by directly comparing bacterial relative abundances with immune module ssGSEA scores across participants. These analyses confirmed the negative associations between the abundances of *C. accolens*, *C. tuberculostearicum*, and *S. epidermidis* and innate immune and inflammation module scores, with *S. aureus* abundance showing positive associations with these same modules (**Fig. 3E**). Together, these findings identify a coordinated baseline nasal microbiome–host immune state associated with susceptibility to SARS-CoV-2 infection.

Similar analyses of throat samples identified specific microbe–host immune response relationships associated with subsequent infection. At baseline (**Fig. S5C**), the relative abundances of *G. haemolysans*, *S. oris*, and *S. pneumoniae* were positively correlated with the expression of immune modules associated with infection, corresponding to ISGs (M3.4), TLR signaling (M3.2), innate inflammation (M4.13), phagocytosis (M5.1), and neutrophil migration (M7.1). In contrast, *V. nakazawae* abundance was negatively correlated with expression of these same modules. On the morning after viral inoculation (1am; **Fig. S5D**), positive relationships were observed between *G. haemolysans*, *S. pneumoniae*, and commensal streptococci and modules related to innate immune signaling, whereas *Prevotella* spp., *Schaalia odontolytica*, and *S. oris* were negatively correlated with these modules. Many of these relationships were also observed at subsequent timepoints, with recurring positive associations between streptococci and innate and inflammatory immune modules, although the specific species and immune modules comprising the networks varied over time (**Fig. S5E**). Participant-level analyses further illustrated selected microbe–host immune response relationships, with *S. mitis* abundance positively associated with several innate and inflammatory immune module ssGSEA scores and *V. atypica* and *V. nakazawae* abundances negatively associated with these scores (**Fig. S5F**).

### Gene expression patterns distinguish participants by symptom burden and viral load

Having identified baseline microbiome–host immune signatures associated with infection susceptibility, we next evaluated whether host responses distinguished clinical outcomes among the 18 participants who became infected. We categorized infected participants as having low or high symptom burden based on symptom score AUC, with values below or above the mean defining the low- and high-burden groups, respectively (**Fig. S1**). In baseline nasal samples, overall immune profiles did not differ between participants who developed high versus low symptom burden (PERMANOVA, R²=0.044, p=0.63); however, participants who developed a high symptom burden had significantly lower expression of immune modules corresponding to monocyte/APC activation (M4.6) and neutrophil degranulation (M5.7) at baseline (**Fig. 4A**). Comparing immune module expression after viral inoculation to baseline (**Fig. 4B**), participants with low symptom burden showed a pattern of early upregulation of multiple immune modules, including modules related to interferon responses, innate inflammation, monocyte/APC activation, neutrophil responses, and B cells, although changes in the expression of individual modules were not statistically significant. Interferon responses (M1.2, M3.4) were also upregulated at later timepoints in this group. Participants who developed a high symptom burden had comparatively less immune activation during this early period, but showed significant upregulation of neutrophil degranulation (M5.7) by 2am, followed by more pronounced upregulation of interferon responses and modules for monocyte/APC activation (M4.6) and NK cells (M3.6) (**Fig. 4B**). We next directly compared expression of selected immune modules over time between participants with low and high symptom burden (**Fig. 4C**). Participants who developed high symptom burden had lower expression of several of these modules at baseline but rapidly increased their expression following inoculation, reaching levels comparable to those in participants who developed low symptom burden. Monocyte/APC activation (M4.6) and neutrophil degranulation (M5.7) subsequently increased more sharply and were generally higher in participants with high symptom burden, while type I interferon signaling (M1.2) showed a more rapid increase than in participants with low symptom burden (**Fig. 4C**). Together, these findings suggest that lower symptom burden among SARS-CoV-2-infected individuals is associated with earlier nasal immune activation following viral exposure, whereas higher symptom burden is characterized by lower baseline nasal expression of selected innate immune modules and more pronounced immune activation later in infection.

**Figure 4.**
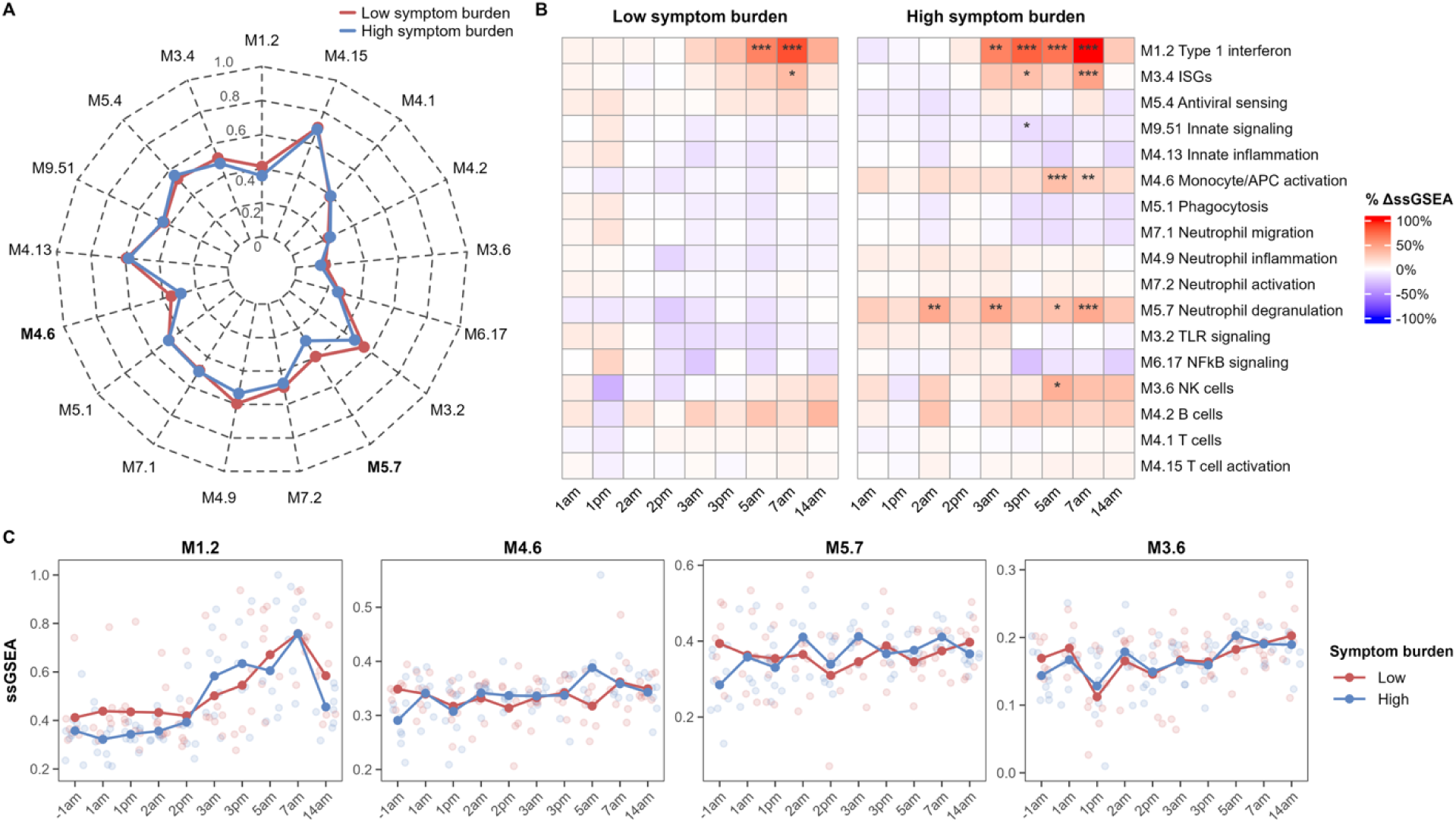
Nasal immune module expression by symptom burden among infected participants. **(A)** Baseline (−1am) immune module enrichment scores in infected participants stratified by symptom burden. Radar plots display mean scaled ssGSEA values for participants with low (red) and high (blue) symptom burden. Modules with baseline differences at q < 0.20 based on age- and sex-adjusted linear regression analyses are shown in bold. Scaled ssGSEA values were normalized to a 0–1 range within the nasal dataset. **(B)** Longitudinal changes in immune module expression among infected participants with low or high symptom burden. Heatmaps show percent change in mean scaled ssGSEA values from baseline, with red indicating increased expression and blue indicating decreased expression. Asterisks denote significant within-group changes from baseline estimated using linear mixed-effects models adjusted for age and sex with participant-level random effects (*q < 0.20, **q < 0.05, ***q < 0.01). **(C)** Longitudinal trajectories of selected immune modules associated with symptom burden among infected participants. Points represent individual participant observations and lines indicate group mean scaled ssGSEA values over time.

In contrast to nasal samples, there were no significant differences in baseline immune module expression in throat samples from participants with high versus low symptom burden (**Fig. S6A**). Starting on day 3, both high and low symptom burden participants upregulated modules related to interferon responses (M1.2, M3.4) and antiviral sensing (M5.4) (**Fig. S6B**). Notably, participants with low symptom burden exhibited broader immune activation in the throat, with upregulation of modules related to innate inflammation and signaling (M4.13, M9.51), phagocytosis (M5.1), and neutrophil migration (M7.1). Direct comparison of module expression over time showed similar induction of interferon responses (M1.2, M3.4) in both groups, whereas expression of innate inflammation (M4.13) and phagocytosis (M5.1) modules were generally higher in participants with low symptom burden following their induction at the 3pm timepoint (**Fig. S6C**).

We conducted similar analyses comparing gene expression in nasal and throat samples from participants with high versus low viral load, with cumulative viral load AUC values below or above the median defining the low- and high-viral-load groups, respectively (**Fig. S1**). There were no baseline differences in immune module expression by viral load in nasal samples (PERMANOVA, R²=0.074, p=0.35, **Fig. S7A**). After inoculation, both groups showed strong upregulation of interferon responses, although this response tended to occur earlier in participants with high viral loads than in those with low viral loads (**Fig. S7B**). Participants with high viral loads also showed upregulation of modules corresponding to neutrophil degranulation (M5.7) and B cells (M4.2), and downregulation of innate immune signaling (M9.51), beginning around 3am. In throat samples, there were no baseline differences in immune module expression by viral load (PERMANOVA, R²=0.037, p=0.75, **Fig. S7C**). Similar to nasal samples, both groups showed upregulation of modules corresponding to interferon responses and antiviral sensing starting at 3pm. Other immune modules showed comparatively modest changes, with generally similar patterns between participants with high and low viral loads (**Fig. S7D**). Thus, viral load kinetics are associated with the timing of nasal immune responses, whereas immune responses in the throat were broadly similar between viral load groups.

### Microbiome-host immune response networks are associated with symptom burden and viral load

Using DIABLO, we similarly identified microbiome and host immune features associated with symptom burden in nasal and throat samples from participants who developed SARS-CoV-2 infection. In models using baseline (−1am) nasal samples to predict development of low versus high symptom burden, the microbiome and host immune data showed strong discriminatory performance, with AUCs of 0.98 and 0.86, respectively, although classification accuracy was poor (52%) (**Fig. 5A**). AUCs remained >0.70 for both data types at nearly all subsequent timepoints, while classification accuracy peaked at 62% on day 2 after inoculation. At baseline, higher expression of monocyte/APC activation (M4.6), neutrophil inflammation and degranulation (M4.9, M5.7), TLR signaling (M6.17), and NK cell (M3.6) modules, together with higher relative abundances of *C. propinquum*, *F. necrophorum*, and *S. aureus*, were associated with subsequent low symptom burden (**Fig. 5B**). In contrast, higher relative abundances of *C. accolens* and *C. pseudodiphtheriticum* were associated with subsequent high symptom burden. The relative abundances of these species were strongly negatively correlated with the expression of modules for ISGs, monocyte/APC activation, and TLR signaling, whereas species associated with low symptom burden showed opposing relationships with these modules (**Fig. 5C**). Later in the course of infection, higher relative abundances of *F. necrophorum*, *S. aureus*, and *S. epidermidis* were positively correlated with multiple immune response modules, together forming a network associated with low symptom burden (**Fig. 5B, 5D**).

**Figure 5.**
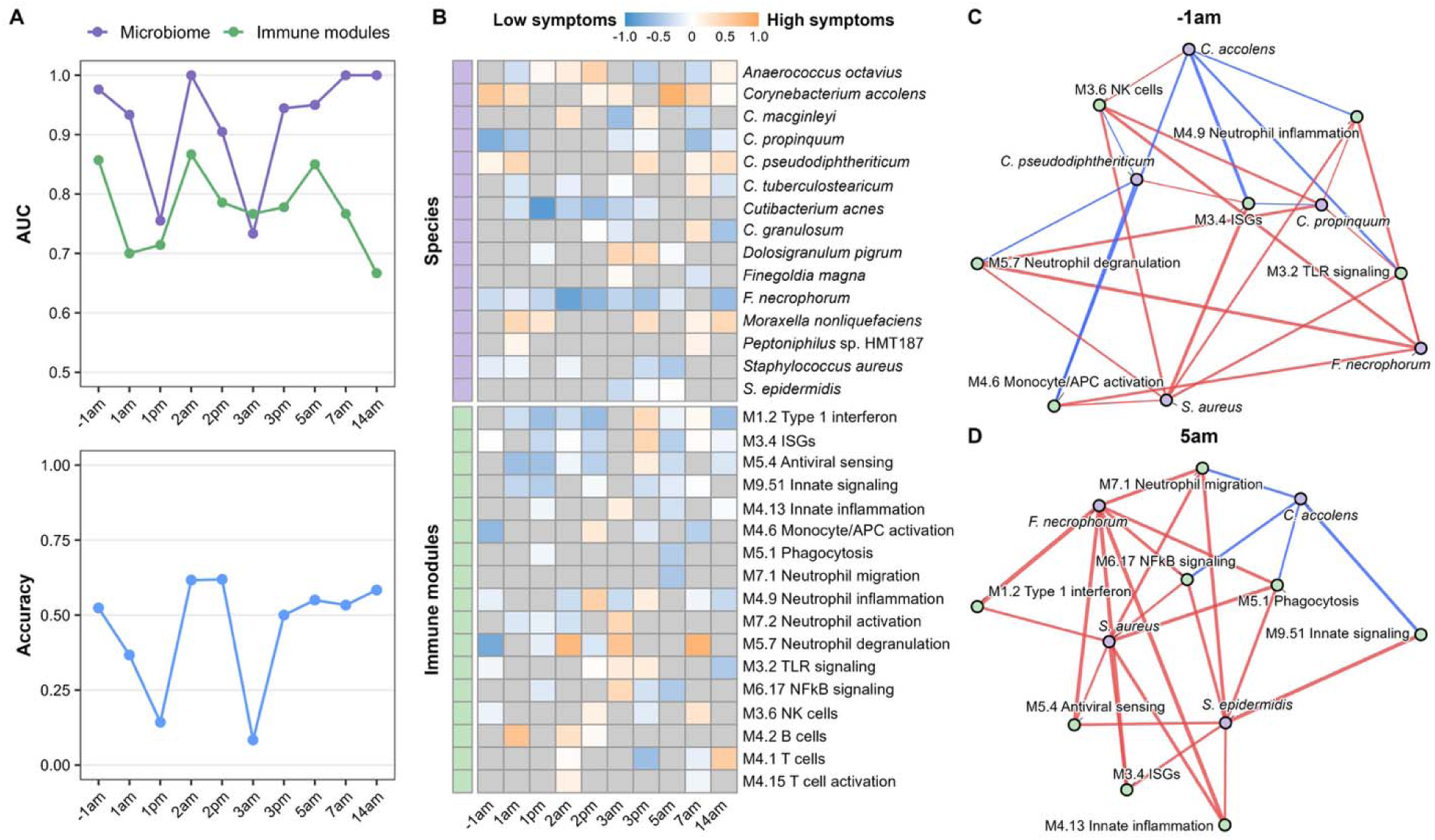
Nasal microbiome-host immune signatures associated with symptom burden among SARS-CoV-2-infected participants. **(A)** Performance of DIABLO models trained to distinguish infected participants with high versus low symptom burden at each study timepoint using nasal bacterial species abundance and host immune module expression data. The upper panel shows component 1 area under the receiver operating characteristic curve (AUC) for microbiome and immune-module data blocks. The lower panel shows overall classification accuracy based on Mahalanobis distance. **(B)** Heatmap of signed component 1 loadings from DIABLO models across longitudinal timepoints. Loadings were oriented such that positive values indicate features associated with high symptom burden and negative values indicate features associated with low symptom burden. Species and immune modules selected by DIABLO are shown. **(C, D)** Microbe–host immune networks at baseline (−1am) and 5am. Networks were constructed using DIABLO-selected microbial species and immune modules. Edges represent Pearson correlations between log_10_-transformed microbial relative abundance and scaled ssGSEA immune-module scores, with edge color indicating positive (red) or negative (blue) associations and edge width indicating correlation strength. To enhance interpretability, only the 25 strongest correlations at each timepoint are displayed.

Models built with data from throat samples showed generally strong discriminatory performance for both microbiome and immune module data across the time course, with AUCs ≥0.70 at most timepoints for both data types, peaking at 0.95 for immune modules at 1pm and 0.93 for the microbiome at 2pm (**Fig. S8A**). In contrast, classification accuracy was highly variable, ranging from 26% to 70%, with only 33% accuracy at baseline (−1am) in predicting whether participants would subsequently develop low or high symptom burden. In contrast to the nasal models, in which higher baseline expression of several innate immune modules was associated with subsequent low symptom burden, higher expression of multiple innate and adaptive immune modules in the throat was associated with high symptom burden (**Fig. S8B**). However, this relationship shifted over the course of infection, with higher expression of many of these same immune response modules associated with low symptom burden at later timepoints. Throat microbiome features associated with symptom burden showed less consistent relationships across timepoints and less evidence of coordinated associations with host immune features than those observed in nasal samples.

Finally, we evaluated associations between microbiome composition, host immune responses, and viral load. We categorized infected participants as having low or high viral load based on cumulative viral load AUC (**Fig. S1**). In nasal samples, models using both microbiome and immune module data showed relatively high AUCs at early timepoints, with classification accuracy reaching ∼75% at 1am (**Fig. S9A**). At this timepoint, higher expression of type I interferon (M1.2), phagocytosis (M5.1), neutrophil-related modules (M4.9, M5.7, M7.2), and NK cells (M3.6) was associated with low viral load, whereas higher expression of modules for monocyte/APC activation (M4.6), B cells (M4.2), and T cell activation (M4.15) was associated with high viral load (**Fig. S9B**). Among microbiome features, higher relative abundances of *C. propinquum* and *D. pigrum* were consistently associated with low viral load across timepoints, whereas higher relative abundances of *Peptoniphilus* sp. HMT187 and *S. aureus* were associated with high viral load. At 1am, when model discriminatory performance was greatest, *C. propinquum* and *D. pigrum* were positively correlated with type I interferon (M1.2), neutrophil-related (M4.9, M5.7, M7.2), and phagocytosis (M5.1) modules, whereas *Peptoniphilus* sp. HMT187 was negatively correlated with many of these same modules (**Fig. S9C**). At 7am, around the time of peak viral load (**Fig. S1**), many of the same *D. pigrum*–immune module correlations persisted, while *C. acnes* was associated with high viral load at this timepoint and showed strong negative correlations with many of these same immune modules. In throat samples, models using both microbiome and immune module data also showed generally strong discriminatory performance by AUC, with AUCs ≥0.70 at most timepoints for both data types; classification accuracy was highly variable across timepoints (13%–77%) (**Fig. S10A**). Multiple immune modules were associated with low viral load in throat samples, including several modules corresponding to innate immune responses (**Fig. S10B**). Several bacterial species also showed associations with viral load across timepoints, including *Veillonella rogosae*, which was associated with low viral load, and *Prevotella veroralis* and *S. oris*, which were associated with high viral load.

## DISCUSSION

Prior studies identified associations between the URT microbiome and respiratory viral infection susceptibility and severity, but our understanding of how specific microbes interact with the host mucosa to shape immune tone and influence infection outcomes remains limited. Moreover, most studies evaluating the URT microbiome and host responses during respiratory infections have been cross-sectional, limiting our ability to distinguish features that precede infection from those that emerge following infection. To address this gap, we leveraged samples and data from a first-in-human SARS-CoV-2 challenge model (*16*). Using bulk RNA sequencing and microbiome profiling of nasal and throat swab samples, we identified URT microbiome features and host immune responses associated with susceptibility to SARS-CoV-2 infection, as well as symptom burden and viral load among infected participants. Further, integrated analyses identified networks of microbiome features and host immune modules that were associated with susceptibility to infection, symptom burden, and viral load, identifying potential microbe–host interactions that may contribute to these outcomes.

The URT is the site of the first interactions between respiratory viral pathogens and the human host; thus, initial host responses within the URT play a critical role in infection susceptibility. We found that baseline nasal immune tone differed between participants who went on to develop infection and those who remained uninfected, with those who became infected exhibiting greater activation of modules for interferon and innate immune responses before viral inoculation. Habibi et al. similarly found that baseline neutrophilic inflammation signatures were associated with the development of symptomatic infection after challenge in an RSV human challenge study (*19*). We also identified differential immune responses by infection status shortly after viral inoculation. Despite lower baseline expression of several immune response modules, uninfected participants mounted a rapid transcriptional response following inoculation, whereas infected participants exhibited a delayed response. Similarly, single-cell multi-omic profiling of samples from the present cohort revealed that immune cells infiltrated the URT in both uninfected and infected participants; however, infiltration occurred earlier and more transiently in uninfected participants (1am) than in infected participants (5am) (*20*). Further, immune cell infiltration was associated with upregulation of interferon and innate inflammatory pathways and neutrophil activation. Previous studies of natural SARS-CoV-2 infection have also identified differences in the magnitude and kinetics of antiviral responses associated with infection and disease severity, particularly with regard to early interferon responses (*21, 22*). SARS-CoV-2 infection can suppress host interferon responses, and individuals who develop severe COVID-19 are more likely to harbor defects in interferon signaling components (*23, 24*). Cheemarla et al. used a nasal organoid model to demonstrate that rapid induction of ISGs prevented SARS-CoV-2 replication, while blocking ISG induction enhanced viral replication (*21*). Similarly, Hatton and colleagues showed that pre-treatment of human nasal epithelial cell cultures with type I and II interferons enhanced antiviral responses and reduced SARS-CoV-2 replication (*25*). In our study, participants who remained uninfected had lower baseline expression of modules corresponding to interferon and innate immune responses those who became infected, but rapidly upregulated these responses within 24–36 hours of inoculation. Taken together, these findings demonstrate that the kinetics of the host response plays a pivotal role in protection against SARS-CoV-2 infection.

Previous studies identified associations between URT microbiome features and outcomes following exposure to respiratory viruses. Using respiratory samples from contacts in an influenza household transmission study, Tsang et al. reported that individuals with greater abundances of *Streptococcus* spp. and *Prevotella salivae* were less susceptible to influenza A virus (*5*). Similarly, Lee and colleagues found that nasal/oropharyngeal microbiome community state type and the abundances of *Alloprevotella*, *Prevotella*, and *Bacteroides* were associated with infection among household contacts of individuals with influenza (*26*). In contrast, in a respiratory syncytial virus challenge study, no differences in URT microbiome composition were identified at baseline between participants who developed infection and those who remained uninfected, nor among infected participants according to symptom burden or viral load (*27*). Several prior studies identified differences in the abundance of specific bacterial species, including *Corynebacterium*, *Dolosigranulum*, *Prevotella*, and *Veillonella* spp., were associated with SARS-CoV-2 infection susceptibility and severity (*11, 12, 28, 29*). However, as is common in studies of natural infection, these studies relied on samples acquired after viral exposure, limiting their ability to identify features present before infection that may be associated with subsequent SARS-CoV-2 infection. In the present study, participants who subsequently became infected tended to have lower relative abundances of *C. accolens* and higher relative abundances of *D. pigrum* and *S. aureus* at baseline, although these differences did not reach statistical significance. In contrast, baseline throat microbiome composition was highly similar between these groups. Together, these findings suggest that the nasal microbiome is associated with SARS-CoV-2 infection susceptibility and highlight specific microbial features for further investigation in larger studies of SARS-CoV-2-exposed individuals.

Microbes in the URT can modify host mucosal immune responses to prevent or promote respiratory viral infections (*6, 7*). This has been demonstrated in human epithelial cell models, where exposure to *Haemophilus influenzae*, *Pseudomonas aeruginosa*, and *Streptococcus pneumoniae* has been shown to alter epithelial responses to subsequent respiratory viral infection and enhance virus-induced inflammation (*8, 10*). In animal models, *S. aureus* and *S. pneumoniae* secrete proteases that cleave influenza hemagglutinin, enhancing viral infectivity, while *S. pneumoniae* reduces adaptive immune responses and directly modifies the host mucosa, potentiating viral infections (*9, 30, 31*). Conversely, the presence of the nasal commensals *Rothia mucilaginosa* and *S. epidermidis* enhanced protection against influenza A virus infection in mice, increasing viral clearance and reducing inflammatory gene expression and lung damage (*32, 33*). Together, these studies demonstrate diverse direct and indirect interactions between URT microbes, respiratory viruses, and host immunity that may influence respiratory viral infection outcomes. Importantly, most studies evaluating the mechanisms by which microbes influence host responses in the URT have been conducted in animal models and have focused on a single bacterial species, limiting their ability to capture the complexity of the URT microbiome. Moreover, differences in host biology between animals and humans further limit the extent to which findings from these studies are likely to reflect microbe–host interactions in humans. Integrated multi-omics approaches that account for multiple microbiome features together with host responses offer an opportunity to identify such associations that can then be further explored in mechanistic studies (*34, 35*). To this end, we used such an approach to characterize coordinated networks linking microbiome features with host immune responses associated with SARS-CoV-2 infection susceptibility, symptom burden, and viral load. These findings provide new insight into how the URT microbiome may influence the local mucosal immune environment, which, together with the microbial community itself, may influence outcomes of respiratory viral infections.

Our study has several strengths and limitations. The human challenge model provided a defined viral inoculum and precisely timed exposure, enabling repeated, dense sampling of a well-characterized cohort. We additionally characterized both host gene expression and microbiome composition within the same samples, permitting an analysis of the relationships between resident microbes and host responses. Limitations include the small number of participants, which limited statistical power for analyses of high-dimensional microbiome and host gene expression data, as well as the inclusion of healthy young adults who generally experienced mild illness. Further, we used bulk RNA sequencing to evaluate host gene expression, limiting our ability to identify specific cell types involved in the response to viral inoculation. Further studies using higher-resolution approaches to characterize immune responses in cohorts spanning different ages, health statuses, and disease severities could provide further insights into the relationship between the URT microbiome and host responses to SARS-CoV-2.

## Conclusions and future studies

Using data from a first-in-human challenge study, we identified coordinated microbiome-host immune signatures associated with SARS-CoV-2 infection susceptibility, symptom burden, and viral load. We identified differences in the kinetics of the early host response associated with infection susceptibility, with a more rapid interferon-based response observed among participants who remained uninfected. Further, we identified URT microbiome features associated with host mucosal immune responses both at baseline and following viral inoculation, suggesting that microbes in the URT contribute to the local immune environment and the host response to viral infection. Taken together, our findings indicate that the URT microbiome and local host immune environment are associated with SARS-CoV-2 susceptibility and begin to define how interactions between these components of the URT ecosystem may contribute to differences in the clinical course of infection. Further studies exploring the mechanisms underlying these relationships hold promise for advancing our understanding of how the URT microbiome shapes mucosal host immunity and informing the development of microbiome-targeted therapies for the prevention or treatment of respiratory infections.

## MATERIALS AND METHODS

### Study design and population

All data and samples used in this study were previously collected through a SARS-CoV-2 human challenge study (ClinicalTrials.gov identifier: NCT04865237). These data and samples were fully de-identified prior to transfer to Duke University for the proposed research. A protocol describing the proposed secondary analyses of these data and samples was reviewed by the Duke University Health System (DUHS) Institutional Review Board and determined to meet the definition of research not involving human subjects as described in 45 CFR 46.102(f), 21 CFR 56.102(e) and 21 CFR 812.3(p). Briefly, healthy adults 18–30 years of age without history of prior SARS-CoV-2 infection or vaccination were recruited to this single-center, phase 1, open-label, first-in-human study conducted between March and July 2021 (*16*). Prior to inoculation, volunteers underwent screening with the BioFire® FilmArray Respiratory Panel 2.1 (BioMérieux) and were excluded if any pathogen was identified. Participants were intranasally inoculated with 10 TCID_50_ of wild-type SARS-CoV-2 (B.1 lineage, D614G variant). Symptom diaries were completed three times daily from day −1 (baseline) through day 14 after inoculation, recording 19 upper respiratory, lower respiratory, and systemic symptoms on a 0–3 severity scale (0 = none, 1 = mild, 2 = moderate, 3 = severe). Participants remained in a high-containment quarantine unit for a minimum of 14 days, with discharge criteria based on virological testing of respiratory samples.

### Host gene expression and microbiome sequencing

Mid-turbinate nasal and throat swabs were collected twice daily, placed in viral transport medium (Bio-Serv), and stored at −80 °C. Samples were analyzed for SARS-CoV-2 by quantitative RT-PCR to determine viral load and by quantitative culture to detect replication-competent virus. For the analyses presented herein, nasal and throat samples collected on day –1 (baseline), twice daily from days +1 to +3 after inoculation, and on days +5, +7, and +14 underwent RNA sequencing for host gene expression profiling and DNA sequencing for microbiome characterization. RNA was extracted using QIAamp Viral RNA kits (Qiagen). Libraries were prepared with TruSeq RNA Exome kits (Illumina) and sequenced on a NovaSeq X Plus instrument configured for 100 base-pair paired-end sequencing. Raw sequencing reads were assessed for quality using FastQC v0.12.1 and trimmed using Trimmomatic v0.39 (*36, 37*). Median [interquartile range (IQR)] sequencing depth for nasal and throat samples after quality filtering was 69.7 (39.5-119.0) and 122.4 (90.4-164.1) million reads, respectively. The resulting quality-filtered reads were aligned to the human reference genome GRCh38 and a gene count matrix was generated using STAR v2.7.11b (*38*). Gene annotation was performed using GENCODE v48 (*39*), with analyses restricted to protein-coding genes and excluding mitochondrial and ribosomal genes.

DNA was extracted from nasal and throat samples using DNeasy PowerSoil Pro kits (Qiagen), with negative and positive extraction controls included in each batch. Bacterial load was quantified by qPCR targeting the 16S rRNA gene using universal primers 1320F and 1431R and Power SYBR Green PCR Master Mix (Applied Biosystems) on a QuantStudio 6 Pro Real-Time PCR system. A standard curve spanning 10 to 10□16S rRNA gene copies was used for quantification, with samples, standards, and controls run in triplicate. For the throat samples, the extracted DNA underwent library preparation using DNA Prep kits (Illumina), incorporating four negative library controls per batch. Libraries were pooled at equimolar concentrations and sequenced on a NovaSeq X Plus instrument (Illumina) configured for 150-bp paired-end reads. Raw sequencing reads were quality filtered and depleted of host contamination (hg19 reference genome) using default parameters in KneadData v0.12.1. Kraken2 v2.1.3 was used for taxonomic classification and was run in paired-end mode with default parameters against the pre-built PlusPF database (07/14/2025 version), which includes archaeal, bacterial, viral, human, and plasmid sequences (*40*). Species-level abundances were estimated with Bracken v3.0.1, using a database constructed with a read length of 150 bp and the default k-mer length of 35 bp (*41*). Presumed reagent contaminants were identified and removed based on presence in negative control samples or negative correlation with DNA concentration using the frequency method (threshold□=□0.10) implemented in the *decontam* R package v1.26.0 (*42*).

Initial shotgun metagenomic sequencing of nasal samples from four participants yielded 99.94% host DNA, limiting recovery of microbial sequences despite deep sequencing. We therefore characterized the nasal microbiome using V1–V3 16S rRNA gene sequencing with primers 27F and 534R (*43*). Extracted DNA from nasal samples was pooled (∼200 samples per pool) and sequenced on a NextSeq XLeap P1 600-cycle kit (Illumina) configured for 300-bp paired-end reads. Raw sequencing data were processed using DADA2 v1.34.0 to generate amplicon sequence variants (ASVs), which were collapsed to the species level, and taxonomy was assigned using the expanded Human Oral Microbiome Database (*43, 44*). Presumed reagent contaminants were identified and removed based on presence in negative control samples or negative correlation with DNA concentration using the frequency method (threshold = 0.3) implemented in the *decontam* R package v1.26.0 (*42*).

### Statistical analysis

Modules of co-expressed genes with shared functions were defined using the gene sets described by Chaussabel et al., and module expression was quantified by ssGSEA (*45, 46*). Functional annotations for each module were derived using Reactome and Gene Ontology Biological Process enrichment analyses, supplemented by manual review of the genes comprising each module (*47, 48*). We selected 17 modules representing key immune functions observed in the dataset, with final annotations based on these enrichment results and manual review (**Supplemental File 1**). For each anatomical site, count data were normalized using the trimmed mean of M-values method and transformed to log counts per million before ssGSEA. Associations between gene module expression and infection status, symptom burden, or viral load were evaluated using linear mixed-effects models, with participant included as a random effect and the Benjamini–Hochberg correction applied to control the false discovery rate. Cross-sectional analyses were performed at individual timepoints, adjusting for age and sex, as well as for changes relative to baseline where relevant. Microbial α-diversity was assessed using observed features and the Shannon index, while β-diversity was evaluated with Bray–Curtis and Jaccard distances using PERMANOVA, with subject-stratified models applied to account for repeated sampling. For taxonomic analyses, species were retained if present at a mean relative abundance >0.5% and detected in >10% of samples; remaining species were grouped as “Other.” Integration of microbiome and host gene expression data was performed using DIABLO, with separate nasal and throat models constructed at each timepoint to identify correlated microbial species and gene modules associated with infection status, symptom burden, or viral load (*18*). Model parameters, including the number of components and features retained, were selected using repeated cross-validation, with a design parameter of 0.25.

## List of Supplementary Materials

Fig. S1 to S10.

Data file S1

## Data Availability

The RNA sequencing dataset generated in this study is available in the European Genome-phenome Archive (EGA) under accession EGAS50000001440. The shotgun metagenomic and 16S rRNA gene sequencing data used for analyses of the URT microbiome are available in the NCBI Sequence Read Archive under accession PRJNA1160064. De-identified metadata, processed microbial species abundance and gene expression count tables, output files, and scripts used for data analysis are available at https://github.com/mskelly7/SARS2_Challenge.

https://ega-archive.org/studies/EGAS50000001440

https://www.ncbi.nlm.nih.gov/sra/?term=PRJNA1160064

## Acknowledgments

We thank the study participants for their time and commitment. We additionally thank the staff of the Duke Microbiome Center and the Duke Sequencing and Genomic Technologies facility for their assistance in developing methods to analyze these samples. CC is supported by the NIHR Imperial Biomedical Research Centre (BRC) award to Imperial College Healthcare NHS Trust and Imperial College. We thank the following organizations for their invaluable contributions to development and implementation of the SARS-CoV-2 human challenge project: the Royal Free London NHS Foundation Trust, Human Infection Challenge Network for Vaccine Development (HIC-Vac), NIHR Clinical Research Network staff at the Royal Bolton Hospital, and ISARIC4C Investigators (https://isaric4c.net/about/authors/) for providing the clinical material for challenge virus production. The authors gratefully acknowledge support from the UK Vaccine Taskforce of the Department of Business, Energy and Industrial Strategy of Her Majesty’s Government (BEIS) and the Wellcome Trust (grant no. 224530/Z/21/Z) and Kwok Foundation. HW is supported by a HIC-Vac pump-priming fund. The funders had no role in the conceptualization, design, data collection, analysis, decision to publish, or preparation of the manuscript. The views expressed are those of the authors and not necessarily those of the NHS, the NIHR, DHSC or BEIS.

## Author contributions

Conceptualization: JHH, HW, CC, MSK

Methodology: VW, PS, IN, DS, LR, SC, JA, SYK

Investigation: JHH, VW, PS, IN, DS, LR, SC, JA, SYK, CH, KLK, AC, MK, BK, AM, HW, CC, MSK

Analysis: JHH, VW, PS, MSK

Visualization: MSK

Funding acquisition: JHH, MSK

Project administration: JHH, MSK

Supervision: JHH, MSK

Writing – original draft: JHH, MSK

Writing – review & editing: VW, PS, IN, DS, LR, SC, JA, SYK, CH, KLK, AC, MK, BK, AM, HW, CC

