## Supplementary material for "Upper respiratory microbiome and host immune signatures associated with SARS-CoV-2 susceptibility and illness in a human challenge model": Fig. S1 to S10

**Supplemental Materials**

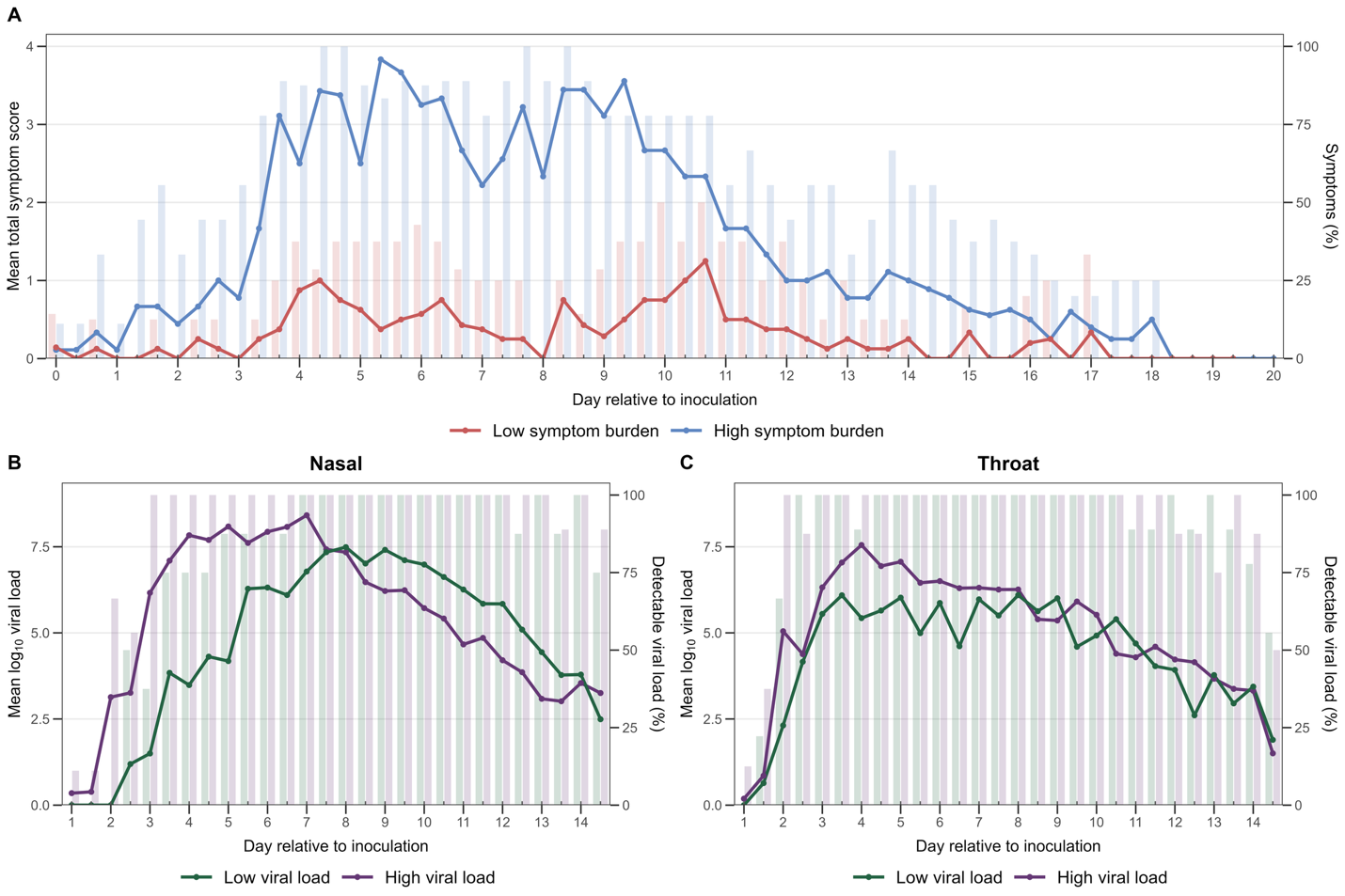
**Figure S1. Longitudinal symptom burden and viral load kinetics following SARS-CoV-2 challenge.** (A) Mean total symptom scores across longitudinal timepoints in participants with low versus high symptom burden. Lines show mean symptom scores at each sampling visit, while bars indicate the percentage of participants with symptoms. (B) Mean log_10_ viral load in nasal samples across longitudinal timepoints, stratified by low versus high viral load. Lines show mean viral load at each sampling visit, while bars indicate the percentage of participants with detectable viral load. (C) Mean log_10_ viral load in throat samples across longitudinal timepoints, stratified by low versus high viral load. Lines show mean viral load at each sampling visit, while bars indicate the percentage of participants with detectable viral load.

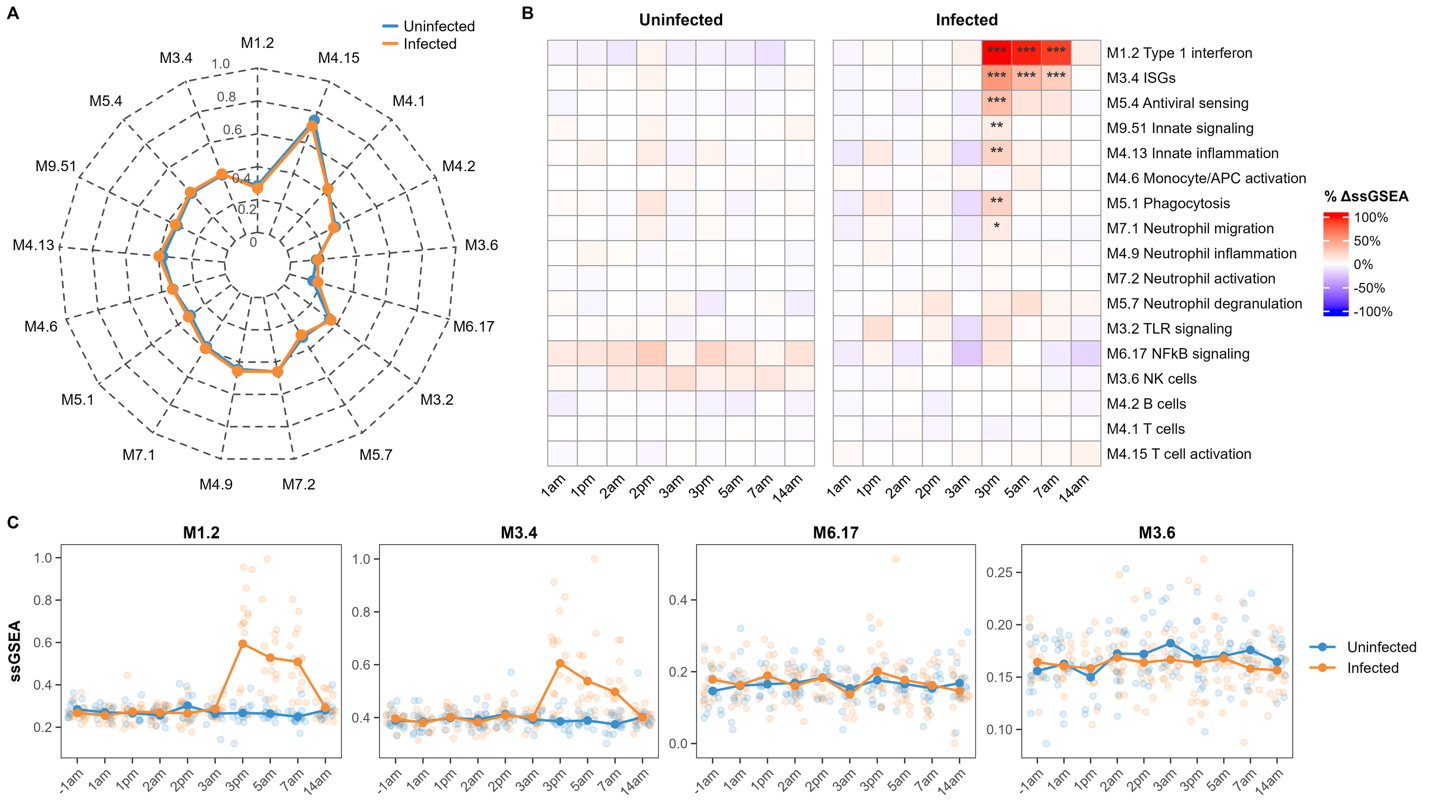
**Figure S2. Throat immune module expression associated with SARS-CoV-2 infection susceptibility. (A)** Radar plot depicting baseline (-1am) throat immune module expression profiles in participants who remained uninfected (blue) or subsequently became infected (orange). Module expression was quantified using single-sample gene set enrichment analysis (ssGSEA) and scaled to a 0–1 range within the throat dataset, where 0 and 1 correspond to the minimum and maximum observed values, respectively. Values represent group mean scaled ssGSEA scores. Module labels shown in bold indicate baseline differences between groups at a false discovery rate (FDR)-adjusted q-value < 0.20. **(B)** Heatmaps depicting longitudinal changes in throat immune module expression relative to baseline within uninfected and infected participants. Values represent percent change from baseline mean scaled ssGSEA scores for each group. Statistical significance was determined using age- and sex-adjusted linear mixed-effects models including timepoint, infection status, and their interaction as fixed effects, with participant included as a random effect. Asterisks indicate FDR-adjusted significance for within-group changes relative to baseline, with FDR correction performed across modules at each timepoint within each infection group (* q < 0.20, ** q < 0.05, *** q < 0.01). **(C)** Longitudinal trajectories of the expression of selected immune modules in throat samples by infection status. Points represent individual participant observations, while lines and larger points represent group mean scaled ssGSEA values at each timepoint.

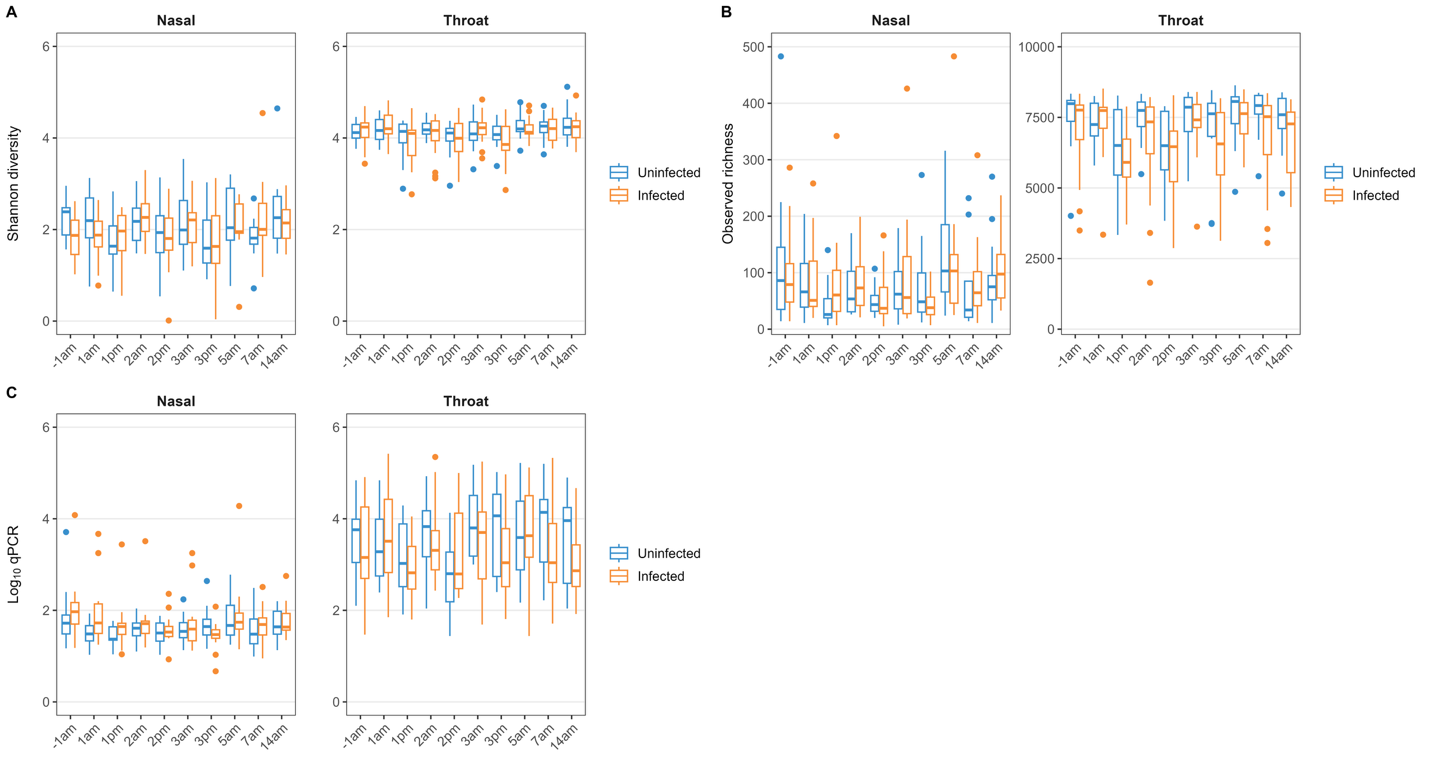
**Figure S3. Longitudinal microbiome alpha diversity and bacterial burden by infection status. (A)** Shannon diversity in nasal (16S rRNA gene sequencing) and throat (metagenomic sequencing) samples across longitudinal timepoints, stratified by infection status. Boxplots display the median and interquartile range (IQR), with whiskers extending to 1.5× the IQR and points indicating outliers. **(B)** Observed richness, defined as the number of detected taxonomic features per sample, in nasal and throat samples across longitudinal timepoints, stratified by infection status. **(C)** Total bacterial burden measured by qPCR and expressed as log_10_-transformed bacterial density across longitudinal timepoints, stratified by infection status. Across all samples, mean bacterial density was approximately 50-fold higher in throat samples than in nasal samples (mean difference, 1.71 log_10_ units). Timepoints are shown relative to viral inoculation, with -1am representing the pre-inoculation baseline sample. Uninfected participants are shown in blue and infected participants in orange.

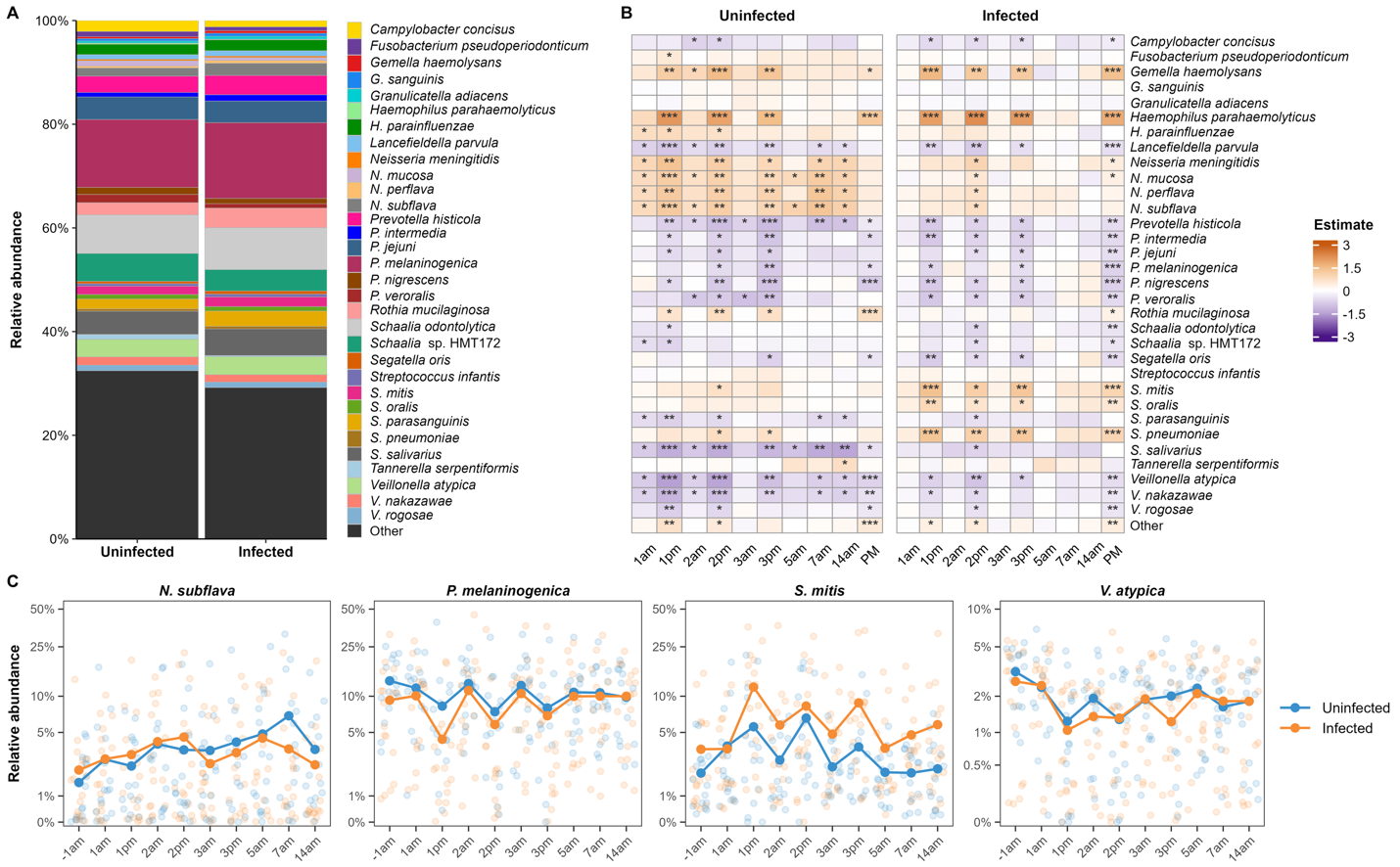
**Figure S4. Throat microbiome composition associated with SARS-CoV-2 infection. (A)** Stacked bar plots depicting baseline (-1am) throat microbiome composition in participants who remained uninfected (blue) or subsequently became infected (orange). Values represent mean relative abundances of retained bacterial species, with remaining species grouped as “Other.” **(B)** Heatmaps depicting longitudinal changes in throat species relative abundance compared to baseline within uninfected and infected participants. Values represent model-estimated changes in centered log-ratio (CLR)-transformed abundance derived from age- and sex-adjusted linear mixed-effects models incorporating day, time of day, infection status, their interactions, and participant-level random effects. Asterisks indicate false discovery rate (FDR)-adjusted significance for within-group changes relative to baseline (* q<0.20, ** q<0.05, *** q<0.01). **(C)** Longitudinal trajectories of selected throat bacterial species by infection status. Points represent individual participant observations, while lines and larger points represent group mean relative abundances at each timepoint. Relative abundance is displayed on a pseudo-logarithmic scale to facilitate visualization of low-abundance species.

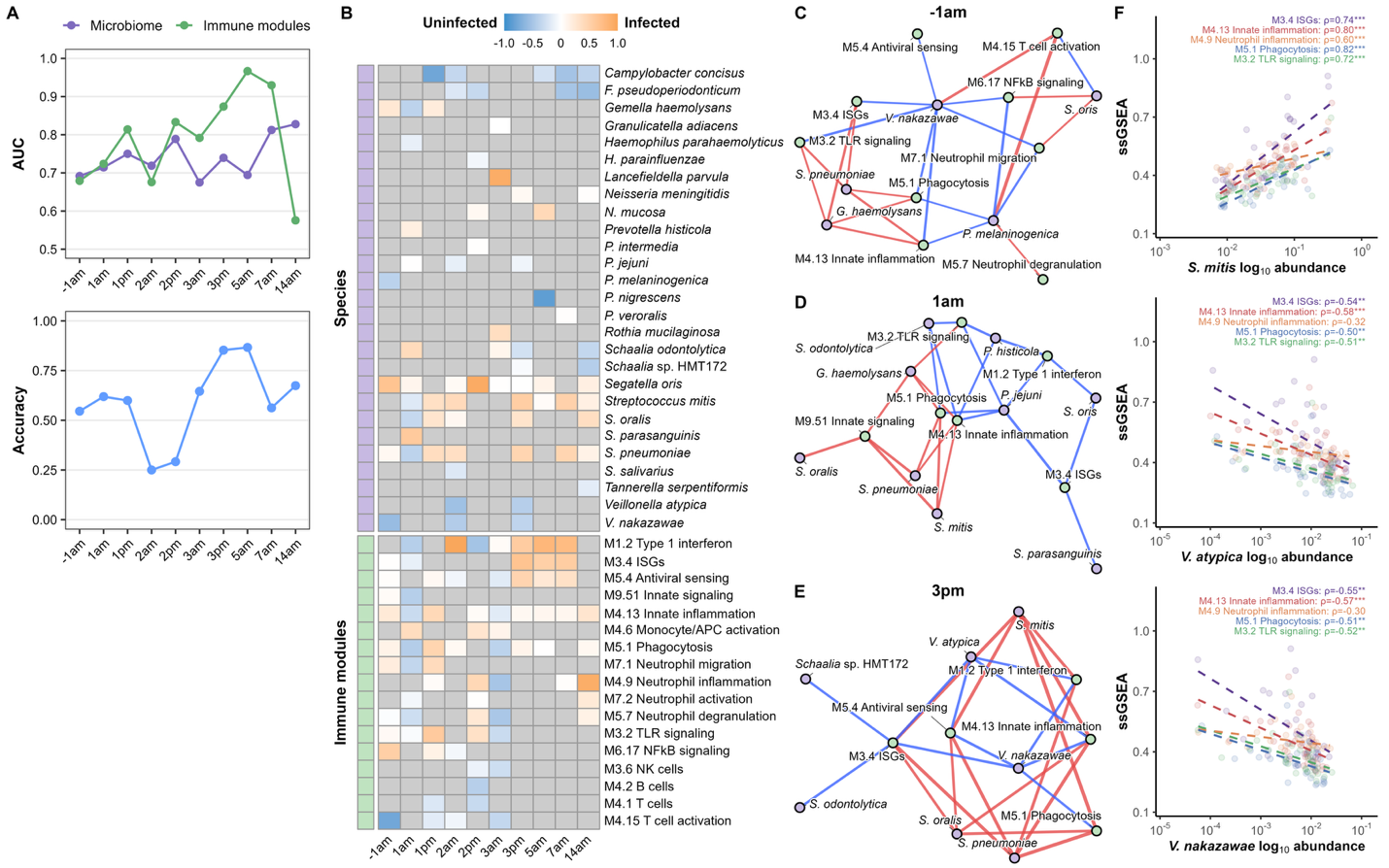
**Figure S5. Throat microbiome-host immune signatures associated with SARS-CoV-2 infection susceptibility. (A)** Performance of DIABLO models trained to distinguish infected and uninfected participants at each study timepoint using throat bacterial species abundance and host immune module expression data. The upper panel shows component 1 area under the receiver operating characteristic curve (AUC) for microbiome and immune-module data blocks. The lower panel shows overall classification accuracy based on Mahalanobis distance. **(B)** Heatmap of signed component 1 loadings from DIABLO models across longitudinal timepoints. Loadings were oriented such that positive values indicate features associated with infected participants and negative values indicate features associated with uninfected participants. Species and immune modules selected by DIABLO are shown. **(C–E)** Microbe–host immune networks at baseline (−1am), 1am, and 3pm. Networks were constructed using DIABLO-selected microbial species and immune modules. Edges represent Pearson correlations between log_10_-transformed microbial relative abundance and scaled ssGSEA immune-module scores, with edge color indicating positive (red) or negative (blue) associations and edge width indicating correlation strength. To enhance interpretability, only the 25 strongest correlations at each timepoint are displayed. **(F)** Associations between selected throat bacterial species and host immune modules at 3pm. Points represent individual participants. Dashed lines represent linear regression fits. Correlation coefficients (ρ) and nominal p values are shown for each immune module.

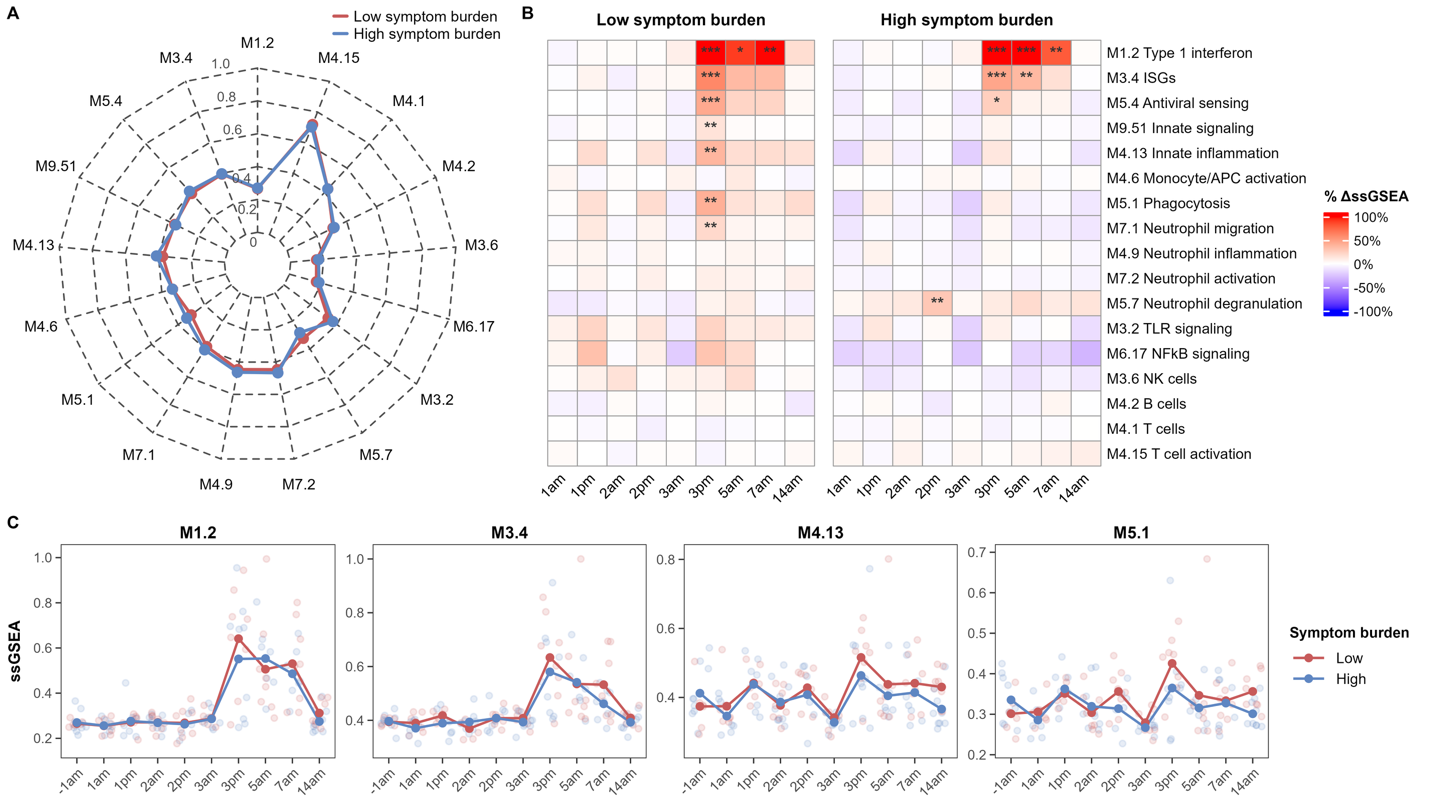
**Figure S6. Throat immune module expression by symptom burden among infected participants. (A)** Baseline (-1am) immune module enrichment scores in infected participants stratified by symptom burden. Radar plots display mean scaled ssGSEA values for participants with low (red) and high (blue) symptom burden. No significant baseline differences were identified in linear regression analyses adjusted for age and sex. Scaled ssGSEA values were normalized to a 0–1 range within the throat dataset. **(B)** Longitudinal changes in immune module expression among infected participants with low or high symptom burden. Heatmaps show percent change in mean scaled ssGSEA values compared with baseline. Red indicates increased expression and blue indicates decreased expression relative to baseline. Asterisks denote significant within-group changes from baseline estimated using linear mixed-effects models adjusted for age and sex with participant-level random effects and false discovery rate correction within each timepoint (*q < 0.20, **q < 0.05, ***q < 0.01). **(C)** Longitudinal trajectories of selected immune modules associated with symptom burden among infected participants. Points represent individual participant observations and lines indicate group mean scaled ssGSEA values over time.

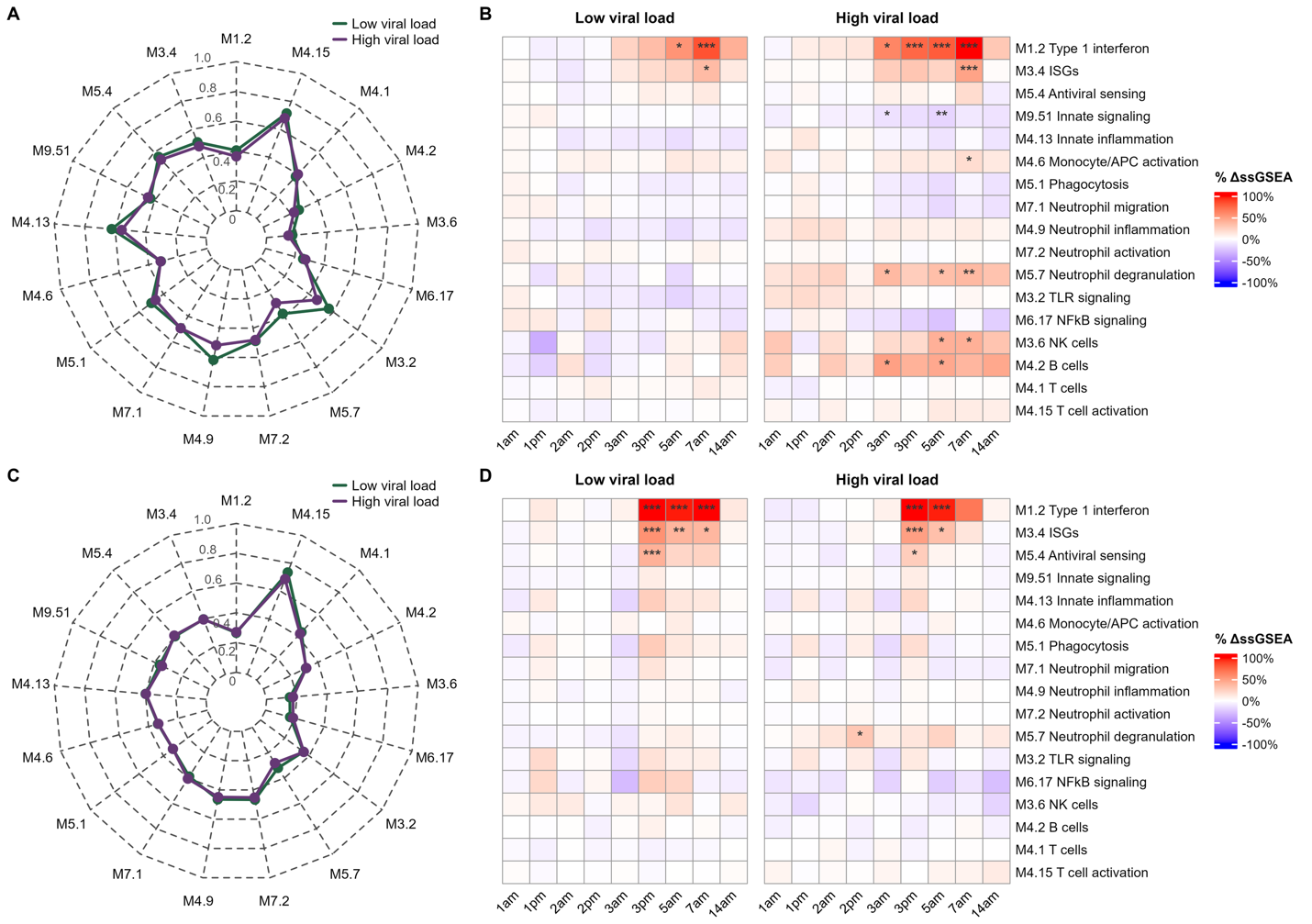
**Figure S7. Nasal and throat immune module expression by viral load among infected participants. (A)** Radar plot depicting baseline (-1am) nasal immune module expression profiles in infected participants with low (green) and high (purple) viral loads. Module expression was quantified using single-sample gene set enrichment analysis (ssGSEA) and scaled to a 0–1 range within the nasal dataset, where 0 and 1 correspond to the minimum and maximum observed values, respectively. Values represent group mean scaled ssGSEA scores. No immune modules demonstrated baseline differences between groups at a false discovery rate (FDR)-adjusted q-value < 0.20. (**B**) Heatmaps depicting longitudinal changes in nasal immune module expression among infected participants with low versus high viral loads. Values represent percent change from baseline mean scaled ssGSEA scores for each group. Statistical significance was determined using age- and sex-adjusted linear mixed-effects models including timepoint, viral load classification, and their interaction as fixed effects, with participant included as a random effect. Asterisks indicate FDR-adjusted significance for within-group changes relative to baseline, with FDR correction performed across modules at each timepoint within each infection group (* q < 0.20, ** q < 0.05, *** q < 0.01). (**C**) Radar plot depicting baseline throat immune module expression profiles in infected participants with low (green) and high (purple) viral loads. Module expression was quantified using ssGSEA and scaled to a 0–1 range within the throat dataset, where 0 and 1 correspond to the minimum and maximum observed values, respectively. Values represent group mean scaled ssGSEA scores. No immune modules demonstrated baseline differences between groups at a false discovery rate (FDR)-adjusted q-value < 0.20. (**D**) Heatmaps depicting longitudinal changes in throat immune module expression relative to baseline within infected participants with low versus high viral loads. Values represent percent change from baseline mean scaled ssGSEA scores for each group. Statistical significance was determined using age- and sex-adjusted linear mixed-effects models including timepoint, viral load classification, and their interaction as fixed effects, with participant included as a random effect. Asterisks indicate FDR-adjusted significance for within-group changes relative to baseline, with FDR correction performed across modules at each timepoint within each infection group (* q < 0.20, ** q < 0.05, *** q < 0.01).

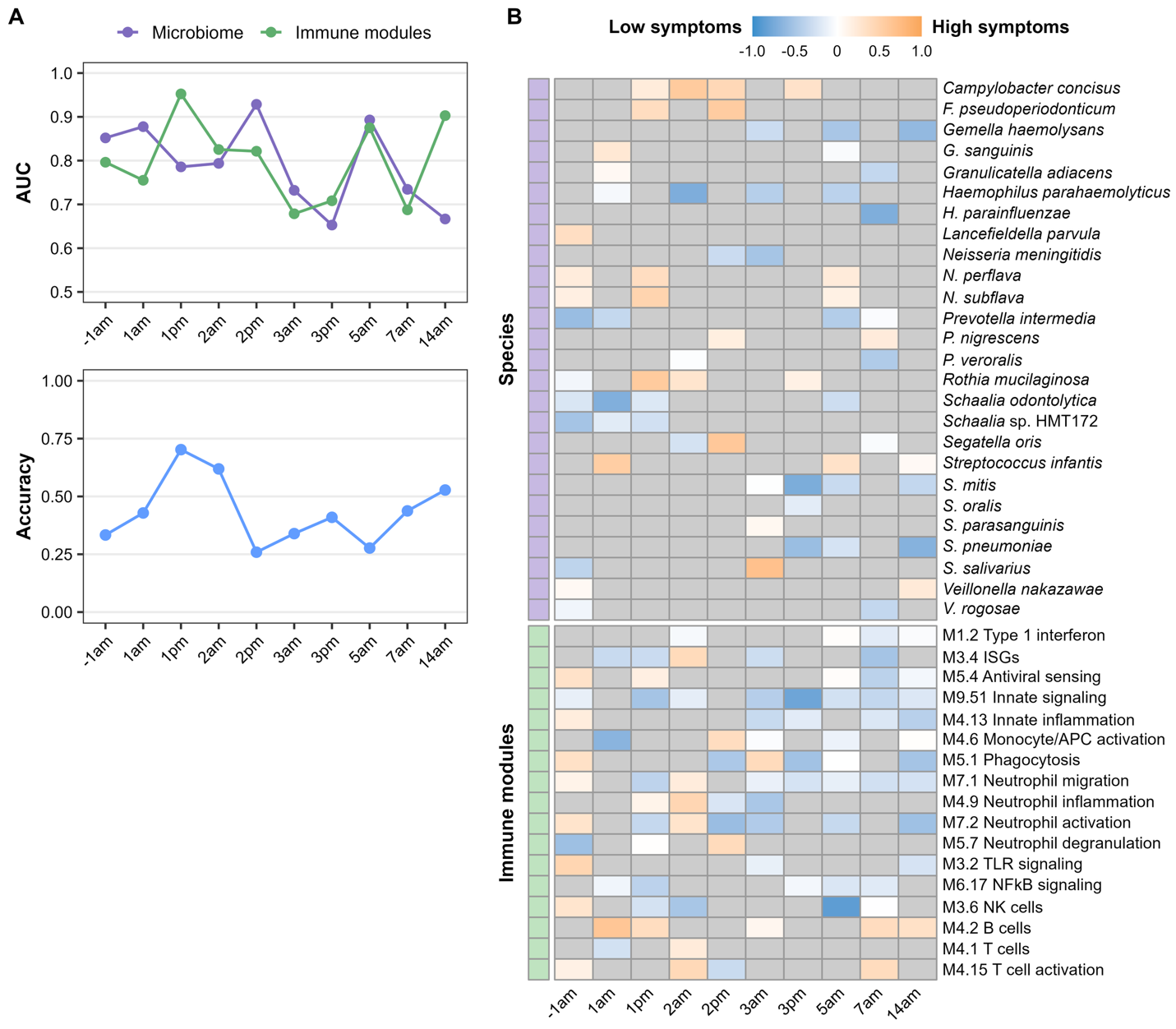
**Figure S8. Throat microbiome-host immune signatures associated with symptom burden among SARS-CoV-2-infected participants. (A)** Performance of DIABLO models trained to distinguish participants with high versus low symptom burden at each study timepoint using throat bacterial species abundance and immune module data. The upper panel shows component 1 area under the receiver operating characteristic curve (AUC) for microbiome and immune-module data blocks. The lower panel shows overall classification accuracy based on Mahalanobis distance. (**B**) Heatmap of signed component 1 loadings from DIABLO models across longitudinal timepoints. Loadings were oriented such that positive values indicate features associated with high symptom burden and negative values indicate features associated with low symptom burden. Species and immune modules selected by DIABLO are shown.

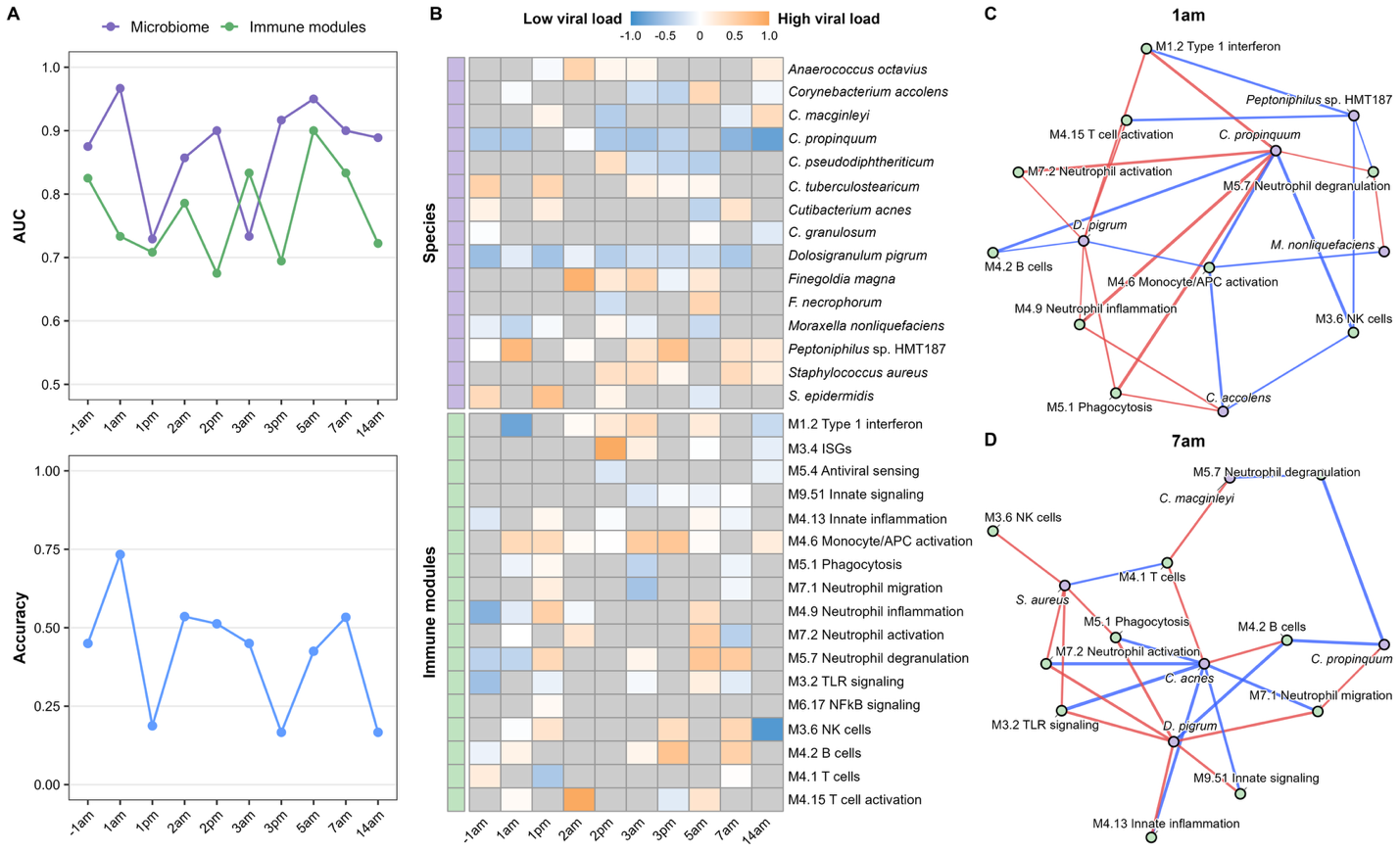
**Figure S9. Nasal microbiome-host immune signatures associated with viral load among SARS-CoV-2-infected participants.** (**A**) Performance of DIABLO models trained to distinguish participants with high versus low viral load at each study timepoint using nasal bacterial species abundance and immune module data. The upper panel shows component 1 area under the receiver operating characteristic curve (AUC) for microbiome and immune-module data blocks. The lower panel shows overall classification accuracy based on Mahalanobis distance. (**B**) Heatmap of signed component 1 loadings from DIABLO models across longitudinal timepoints. Loadings were oriented such that positive values indicate features associated with high viral load and negative values indicate features associated with low viral load. Bacterial species and immune modules selected by DIABLO are shown. (**C**, **D**) Microbe–host immune networks at baseline (−1am) and 7am, with the latter selected to correspond to peak viral load among infected participants. Networks were constructed using DIABLO-selected bacterial species and immune modules. Edges represent Pearson correlations between log_10_-transformed bacterial relative abundance and scaled ssGSEA immune-module scores, with edge color indicating positive (red) or negative (blue) associations and edge width indicating correlation strength. To enhance interpretability, only the 25 strongest correlations at each timepoint are displayed.

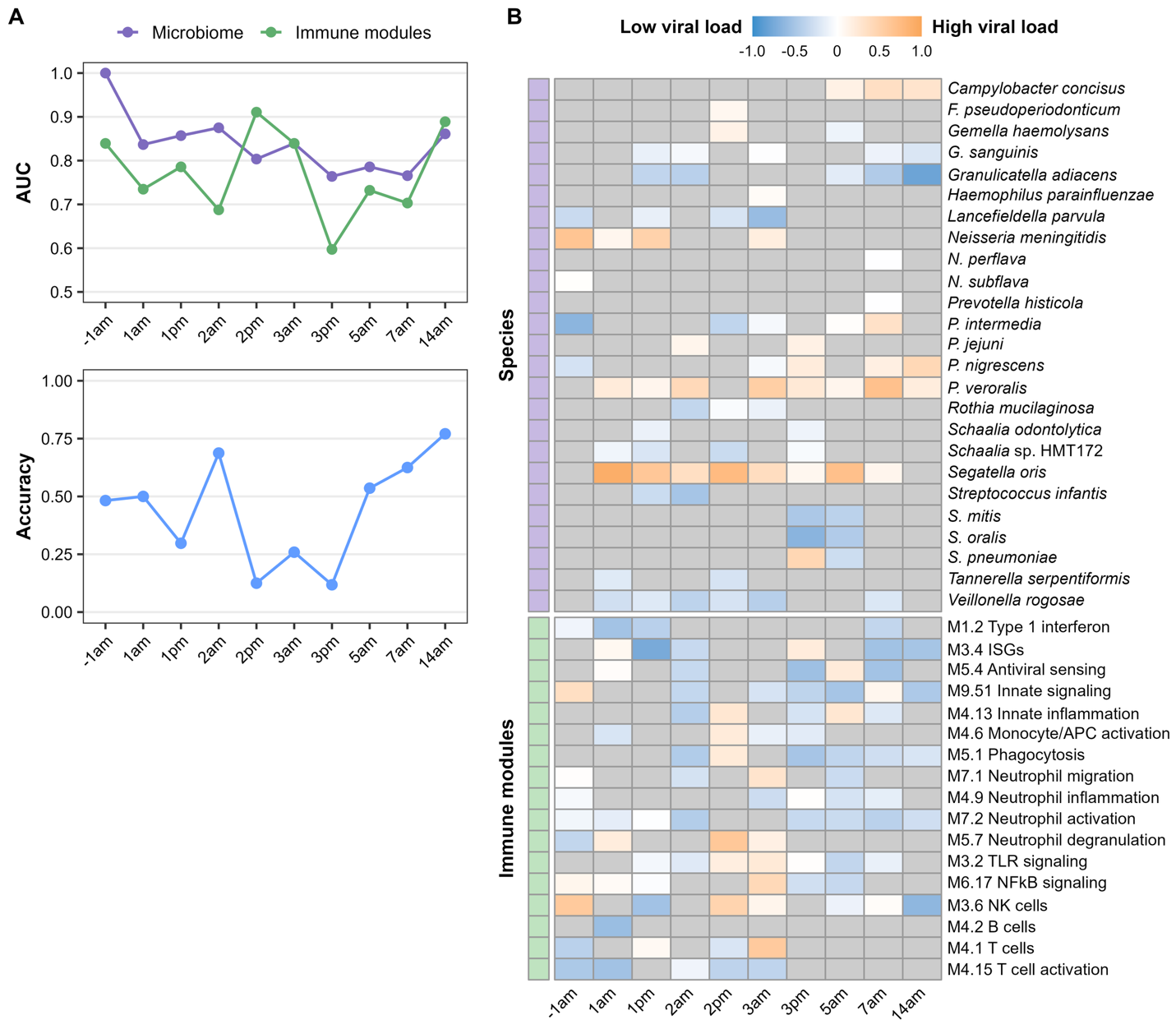
**Figure S10. Throat microbiome-host immune signatures associated with viral load among SARS-CoV-2-infected participants.** (**A**) Performance of DIABLO models trained to distinguish participants with high versus low viral load at each study timepoint using throat bacterial species abundance and immune module data. The upper panel shows component 1 area under the receiver operating characteristic curve (AUC) for microbiome and immune-module data blocks. The lower panel shows overall classification accuracy based on Mahalanobis distance. (**B**) Heatmap of signed component 1 loadings from DIABLO models across longitudinal timepoints. Loadings were oriented such that positive values indicate features associated with high viral load and negative values indicate features associated with low viral load. Species and immune modules selected by DIABLO are shown.
